# Piloting a minimum data set for older people living in care homes in England: a developmental study

**DOI:** 10.1101/2024.06.07.24308589

**Authors:** Adam L Gordon, Stacey Rand, Elizabeth Crellin, Stephen Allan, Freya Tracey, Kaat De Corte, Therese Lloyd, Richard Brine, Rachael E Carroll, Ann-Marie Towers, Jennifer Kirsty Burton, Gizdem Akdur, Barbara Hanratty, Lucy Webster, Sinead Palmer, Liz Jones, Julienne Meyer, Karen Spilsbury, Anne Killett, Arne T Wolters, Guy Peryer, Claire Goodman

## Abstract

**Background:** We developed a prototype minimum data set (MDS) for English care homes, assessing feasibility of extracting data directly from digital care records (DCRs) with linkage to health and social care data.

**Methods:** Through stakeholder development workshops, literature reviews, surveys and public consultation we developed an aspirational MDS. We identified ways to extract this from existing sources including DCRs and routine health and social care datasets. To address gaps we added validated measures of delirium, cognitive impairment, functional independence and Quality of Life to DCR software. Following routine health and social care data linkage to DCRs, we compared variables recorded across multiple data sources, using a hierarchical approach to reduce missingness where appropriate. We reported proportions of missingness, mean and standard deviation (SD) or frequencies (%) for all variables.

**Results:** We recruited 996 residents from 45 care homes in three English Integrated Care Systems. 727 residents had data included in the MDS. Additional data were well completed (<35% missingness at wave 1). Competition for staff time, staff attrition, and software-related implementation issues contributed to missing DCR data. Following data linkage and combining variables where appropriate, missingness was reduced (<=4% where applicable).

**Discussion:** Integration of health and social care is predicated on access to data and interoperability. Despite governance challenges we safely linked care home DCRs to statutory health and social care datasets to create a viable prototype MDS for English care homes. We identified issues around data quality, governance, data plurality and data completion essential to MDS implementation going forward.

**Key points:**

- There is a range of resident information across DCRs, health and social care datasets, which can be combined to provide a more complete picture of residents.
- We developed and implemented a Minimum Dataset linking care home digital care records to statutory health and social care records.
- Information governance for linking data across multiple data owners and data processors is complex and time consuming.
- Standardisation across Digital Care Records Systems would enable data to be used more effectively across the care home sector.
- Establishing shared priorities across key stakeholders interested in care home data is essential for effective MDS implementation.

## Background

Care homes provide around-the-clock residential care for people whose needs cannot be met by visiting care. Older people living in care homes often have needs defined by one or more of frailty, multiple long-term conditions, disability or cognitive impairment [1]. Homes can be registered as with or without nursing depending on whether they employ registered nurses to oversee and provide complex healthcare. In England, there are around 372,000 care home places [2].

Day-to-day care for residents generates abundant data spread across records held by care homes, statutory social care organisations, the National Health Service (NHS), residents and their families [3,4]. As records become increasingly digitised, there is an opportunity to collate data to inform decisions about commissioning, care planning and delivery, review and funding at the micro (individual resident), meso (care home and regional system) and macro (national system) levels [4].

Care home residents were amongst those most adversely affected by COVID-19 and the sector was devastated by outbreaks [5]. At pandemic outset, England lacked even rudimentary data on how many people lived in care homes to track COVID-19 incidence [6]. Emergency legislation, now repealed, enabled collated datasets and recognition of their potential to inform and transform care.

In other countries, Minimum Data Sets (MDSs) for care homes already exist. The most widely recognised of these are the US Medicare Minimum Data Set (MDS 3.0) [7] and InterRAI, deployed in multiple jurisdictions [8]. Implementation of MDSs is influenced by mandates and financial incentives supported by: ongoing training to motivate staff to engage with MDS completion; the extent to which completion is built into the working practices, monitoring, and record systems of all staff (including visiting professionals); and digital recording systems that care home staff use to document and discuss care [9]. At the time of writing, there is no national mandate or incentive framework for implementation of an MDS in any of the four UK nations, although plans are underway to standardise some aspects of social care data collection in England [10].

Against this background, we set out to pilot a prototype MDS for English care homes for older people, focussing on homes currently using digital care records (DCRs) [11]. Our objectives were to: (1) assess feasibility of extracting data directly from DCRs and linking these to routinely collected health and social care data to populate a pilot care home MDS; (2) to assess quality and completeness of MDS data; and (3) describe barriers and facilitators to implementation and use. In this article, we address the first two of these objectives. Implementation and use by care home staff and external stakeholders are addressed in a second paper[12].

## Methods

This was a mixed-methods pilot of a prototype MDS. A full protocol is published elsewhere [11].

### Sampling and resident recruitment

We aimed to recruit 20 care homes for older people in each of three Integrated Care Systems (ICSs), totalling 60 homes. ICSs are regional partnerships between NHS organisations, local government and others including third sector and social enterprises, which are responsible for co-ordinating and commissioning care in England. From the 42 English ICSs we chose three – in the South East, East Midlands, and North East – to sample different geographies, socio-economic deprivation indices, and care configurations. Assuming an occupancy rate of 90%, the sample size required for a true representation of the finite older care home population in each of the ICSs, with 90% confidence and 5% margin of error, was 262-268 residents per ICS [11].

Care homes were eligible for inclusion if using DCRs from one of two participating DCR software companies. Initial approaches were made by email, telephone and in-person, with homes recruited from those responding positively.

All permanent residents of participating care homes were eligible. We excluded: residents receiving respite or temporary/short stay care to minimise burden for people undergoing acute transitions; and residents identified as in the last few days of life by care home staff to protect residents and families at a difficult time. Consent was obtained from residents to access and extract pseudonymised data from their care home, health and social care records and, separately, to link these. Capacity to provide consent to participate was assessed by a researcher at first meeting. For those without capacity, we asked care home staff to send a letter to a family member or friend who could act as a personal consultee as defined by the Mental Capacity Act. Consultee discussions were conducted either face-to-face or by telephone.

### Selecting items for inclusion in the prototype MDS

MDS development was based upon: a review of international research literature summarising outcome measures used in care home studies [13]; a review of measures used in UK care home randomised controlled trials [14]; a systematic review on how contextual factors influence research processes, including data collation in care homes [15]; a series of consultation activities with stakeholders comprising care home managers and staff, and clinical specialists in healthcare of older people and primary care [16–18]; public involvement activity with care home residents, staff and family carers[19]; a survey of data currently collected and collated by English care homes [3]; and a scoping review of published MDSs. From these, we developed nine core principles to govern development and implementation of a care home MDS, previously published [17] (reproduced in Appendix 1).

Based upon these, we compiled an aspirational prototype MDS, containing agreed information and a plan for which routine datasets we hoped to collect these from [11] (summarised in Appendix 2). The systematic review on how contextual factors influence research processes[15] informed our approach to MDS implementation.

### Digital care records (DCRs)

We worked with the Care Software Providers Association (https://caspa.care) to identify two leading care management software providers. Through an initial mapping exercise, based on demonstration of a ‘standard’ user interface by the software providers, we identified variables from the aspirational MDS likely to be included in DCRs.

A dummy data extract from both software providers, completed in summer 2022, identified several variables collected in free text or non-standardised formats. To address gaps in the MDS left by these, that could not be addressed through routine NHS and social care data, additional measures were added to each software system. These included seven validated measures of: delirium (I-AGeD) [19]; cognitive impairment (MDS Cognitive Performance Scale (MDSCPS)) [22]; functional independence (Barthel index)[23]; and Quality of Life (QoL) from the Adult Social Care Outcomes Tool Proxy (ASCOT-Proxy-Resident) [24,25], EuroQol 5 domain 5 level proxy version (EQ-5D-5L Proxy 2) (EuroQol)[26], ICECAP-O [27], and QUALIDEM [28].

The QoL measures were selected based on evidence of use in care homes, psychometric properties [22], relevance to different QoL constructs (health, social care, dementia, and older people), and advice from stakeholder consultations and public involvement activity [18]. Taking into account the high prevalence of cognitive impairment in care home residents[1], proxy versions were used. We further included the ASCOT pain item and low mood/anxiety subscale[30], as well as a question to rate overall QoL on a 7-point scale. This overall question was for resident completion where possible, or otherwise by staff proxy. The type of help needed by the resident, if any, was recorded.

Researchers provided specifications for the user interface format, data extract and outputs for these measures, which were then implemented by software providers and tested by researchers using a pilot interface, with revision as needed. In this process, it became evident that some specifications were not possible in both systems due to differences, for example, in how they dealt with missing data and/or because requirements were incompatible with a system’s usual function or output.

Researchers met with care home staff to describe and explain the additional variables, and to highlight the need for these to be inputted manually in addition to usual care records. For routinely collected variables, data were extracted from existing records without additional input from care home staff, in the format(s) used by care homes and in an output format feasible for each software provider. This minimised burden on care homes and software providers but meant researchers had to clean raw data and derive variables.

All DCR variables were collected twice, six months apart, in March-June and September-November 2023. We collected a small amount of data directly from care homes through a short online survey at baseline to better understand context of care, including number of beds, residents, self-funding residents and staff employed by the care home.

### Routinely collected health and social care datasets

We aimed to access the following data sources: general practice electronic medical records and prescribing data, hospital administrative data, operational datasets from emergency services, urgent care and community health, data from local authorities on social care funding, and data from CQC. We expected to access some of these sources at national (e.g. administrative hospital data) and others at local (e.g. community health) level (Appendix 2).

We developed a data flow diagram (Appendix 3) and legal bases for data sharing (Appendix 4).

### Data management and linkage

As the Improvement Analytics Unit based at The Health Foundation (THF) led data management and linkage, data were hosted on THF’s secure ISO27001/DSPT accredited Data Analysis Platform (DAP). Data were stored in AWS S3 buckets which only Data Managers and approved project data analysts could access. Access to data was controlled by Data Managers.

For extracts of health and social care information held by different data controllers to be created, pseudonymised and shared with THF, we securely transferred to software providers a unique NHS number salt key to enable pseudonymisation of subjects in the study. A separate salt key was used to pseudonymise the Care Quality Commission (CQC) location identifier (unique for each home). Both salt keys used the SHA256 hashing algorithm. Care home pseudonymisation minimised risk of re-identification of individuals based on location. Care home software providers securely transferred extracted DCRs and pseudonymised NHS numbers and care home identifiers for included residents. Data managers isolated pseudonymized NHS numbers and used a pre-computed rainbow table (password cracking tool) of hashed NHS number and salt combinations to determine actual NHS numbers of subjects. These were securely transferred to data processors of health and social care data to enable extraction of relevant records of consented residents. Salt keys were separately transferred so data processors could pseudonymise NHS numbers and care home identifiers in extracted health and social care information. Pseudonymised records were securely transferred to THF once all other identifiers were removed.

Non-personal, aggregated care home-level online survey data from care homes in the study were securely transferred to THF by University of Kent, and pseudonymised by THF.

The salt keys, rainbow table of hashed NHS number and salt combinations, and data from the survey of care providers with clear CQC location identifiers from University of Kent, were stored in a location accessible only by Data Managers, separated from the extracted pseudonymised DCRs and health and social care records, and deleted after the datasets were linked.

Once data were received from data processors, the data was checked and cleaned and variables were derived. Datasets were linked via pseudonymised NHS numbers and pseudonymised CQC location identifiers.

### Stakeholder engagement

We engaged technical experts within NHS England (NHSE) and the ICSs on information governance, data access and availability. We also engaged with wider stakeholders within each ICS to gain support for the project, to facilitate data sharing, and to inform analyses to be conducted on the MDS. Stakeholders included care home managers, staff, residents and family members, GPs, and local decision makers within the NHS and local authority. We also engaged with DHSC and NHSE programme teams (Enhanced Health in Care Homes and Ageing Well) at a national level to understand how an MDS could inform national policy priorities.

Importantly, initial buy-in from the three ICSs at the start of the study, three years before resident consent and data collation began, dissipated by the time discussions around data access started. This was due both to key stakeholders leaving and competing priorities for limited analytical and IG resource. Stakeholders who were able to influence data access and had clinical contact with care homes to inform discussions about data analysis differed between ICSs.

### Deriving MDS variables

We designed a person-level, one row per resident MDS. The date on which additional care home measures were first completed by care home staff, or 1 June if missing, was the index date for all other MDS variables. The Elixhauser list of comorbidities[31,32] and a validated list of frailty syndromes [33] were identified from hospital admission data using ICD-10 codes for 3 years prior to each resident’s index date. Potentially avoidable admissions were those due to a list of conditions originally developed by the Care Quality Commission [34] Healthcare utilisation was collated for the year before the index date. By exception, ambulance activity was only calculated for the period between the first and second MDS measurements. “Out of hours” was defined as 18:00-08:00 and “long attendance” as being at Emergency Departments for 12 hours or longer. All variable derivations are detailed in the final MDS data specification (Appendix 5).

### Data Analysis

Where variables were available from multiple data sources, we compared levels of completeness and agreement. To determine which data source(s) would populate the final MDS, we constructed a hierarchy, based on data quality and expert opinion. We distinguished between variables with a universal definition across datasets, such as date of birth or sex, and those which could be defined in multiple ways or vary over time, such as cognitive impairment or delirium. For the first category, we created a hierarchy collapsing all sources into one final variable. For the second, we presented a comparison but retained all variables in the final MDS. By exception, we took an additive approach for dementia. We used Personal Demographic Service (PDS)[35] as the master index based on NHSE guidance, and Secondary Uses Service (SUS) [36] where data were unavailable in PDS. The exception was ethnicity, where we used the care home record in the first instance, as self-reported ethnicity is more accurate than observational data commonly found in secondary care records [37,38].

Date of death can often generate disagreement between systems, mainly because dates of death notification and certification by the Office of National Statistics may differ [38]. However, they rarely vary more than 30 days, with negligible effect on analysis.

To understand the information contained within the MDS, we reported proportions of missingness, mean and standard deviation (SD) or frequency (%) as appropriate. We also derived two-way tables to provide worked examples of opportunities for more detailed descriptive statistics from the MDS, focussing on emergency attendances and ambulance activity based upon discussions with stakeholders described above.

Evaluation of psychometric properties of the QoL measures (ASCOT-Proxy-Resident, ICECAP-O, EQ- 5D-5L Proxy 2, QUALIDEM) are reported elsewhere [39,40]. These analyses identified limitations around using QUALIDEM in an older adult care home MDS, so we do not report QUALIDEM results here.

The analysis code is published on Github: https://github.com/HFAnalyticsLab. We used R version 4.0.2, SAS Enterprise Guide version 8.3 (NHS and social care routine data), and Stata version 18 (DCR data).

## Results

We recruited 996 residents from 45 care homes (Table Table). Working from lists of care home providers using particular DCR software meant brokering relationships with care homes often new to research. Success was greatest in ICS Area 1 because of long-established relationships between the researchers and their local care home community.

### Digital Care Records (DCRs) from Care Homes

First, we describe DCR data extracted from care homes (Table 2) before we consider accessed datasets and subsequent linkage into the final prototype MDS. Table 2 includes data for the 790 residents (see Figure 1, under consent and extraction) who provided consent and had a valid ID for data extraction (n=748 at Wave 1, n=711 at Wave 2). For residents with complete data at Wave 1, but not Wave 2, most were attributable to resident death or care home drop-out from the study between waves (see Appendix 6).

**Figure 1.**
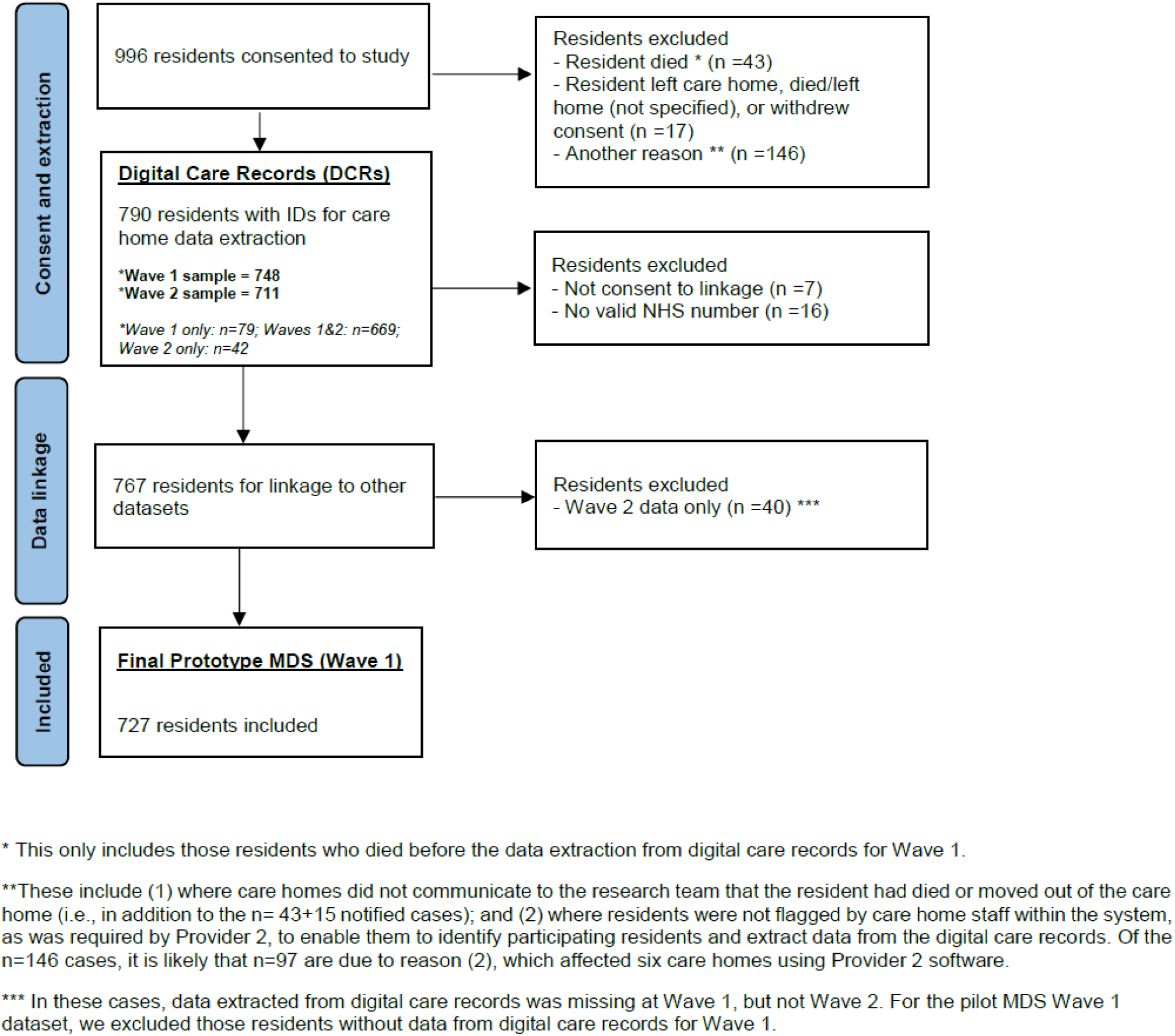
Flow diagram of residents from recruitment into final prototype MDS (Wave 1)

**Table 1.** Actual versus target recruitment by ICS area. From 996 eligible residents, 767 had data extracted which could be linked. Of these, 727 residents had complete data for baseline DCR data collection and were included in the final prototype MDS (Figure 1). Of these, 696 had a DCR with a valid CQC identifier enabling linkage to care home level data from CQC records and the online survey.

| ICS Area | Target recruitment | Actual recruitment |
| --- | --- | --- |
| 1 | 20 care homes | 19 care homes |
|  | 320 residents | 537 residents |
| 2 | 20 care homes | 15 care homes |
|  | 358 residents | 286 residents |
| 3 | 20 care homes | 11 care homes |
|  | 292 residents | 173 residents |
| Total | 60 care homes | 45 care homes |
|  | 970 residents | 996 residents |

**Table 2.** Overview of data extracted from digital care records.

|  |  |  | Wave 1 |  |  | Wave 2 |  |  |  |
| --- | --- | --- | --- | --- | --- | --- | --- | --- | --- |
| MDS | Variable | Categories (if applicable) | n | Mean, SD. (Range) or Freq. % | % Missing data <sup>1</sup> | n | Mean, SD. (Range) or Freq. % | % Missing data <sup>2</sup> | Other comments |
| 1 | Ethnicity | Asian or Asian British<br>White or White British | 163 | ≤4%<br>≥96% | 78.2% | 122 | ≤4%<br>≥96% | 82.8% | Derived variable. Coded differently by the two software providers. |
| 1 | Religion | No religion<br>Christian<br>Buddhist<br>Other | 119 | 11.8%<br>≥76.5%<br>≤5%<br>6.7% | 84.1% | 89 | 9.0%<br>≥79%<br>≤6%<br>≤6% | 87.5% | Derived variable. Coded differently by the two software providers. |
| 1 | Marital Status | Married/cohabit<br>Widowed<br>Divorced/single/separated | 114 | 35.1%<br>50.9%<br>14.0% | 84.8% | 120 | 34.2%<br>49.2%<br>16.6% | 83.1% | Not available for Provider 2 (100% missing). |
| 1 | First Language | English<br>Other | 170 | ≥96%<br>≤4% | 77.3% | 134 | ≥96%<br>≤4% | 81.2% | Not available for Provider 2 (100% missing). |
| 1 | Deprivation of Liberty | No<br>Yes | 689 | 81.3%<br>18.7% | 7.9% | 560 | 76.8%<br>23.2% | 21.2% |  |
| 1 | Weight | 20-35kg<br>36-50kg<br>51-65kg<br>66-80kg<br>81-95kg<br>96-110kg<br>111-125kg<br>126-140kg | 586 | 1.2%<br>19.8%<br>≥39.4%<br>26.3%<br>7.7%<br>3.6%<br>≤1%<br>≤1% | 21.7% | 573 | 1.9%<br>19.2%<br>≥37.6%<br>26.9%<br>9.1%<br>3.3%<br>≤1%<br>≤1% | 19.4% | Derived variable. Provider 1: numerical, Provider 2: categorical. Majority of missing data (≥95%) are from Provider 2. § |
| 1 | Height | 111-125cm<br>126-150cm<br>151-170cm<br>171-190cm<br>191-210cm | 738 | 1.1%<br>9.8%<br>≥70.1%<br>18.0%<br>≤1% | 1.3% | 674 | 1.2%<br>11.0%<br>≥68.2%<br>18.6%<br>≤1% | 5.2% | Derived variable. Provider 1: numerical, Provider 2: categorical. |
| 1 | DNACPR Status | No<br>Yes | 745 | 21.1%<br>78.9% | 0.4% | 707 | 18.7%<br>81.3% | 0.6% |  |
| 1 | Power of Attorney | No<br>Yes | 170 | 62.4%<br>37.6% | 77.3% | 168 | 60.7%<br>39.3% | 76.4% | Not available for Provider 2 (100% missing). |
| 3 | Length of stay, days |  | 747 | 873.7, 807.5 (<50 to >8,000) | 0.1% | 710 | 1,011.4, 815.6 (<50 to >8,000) | 0.1% | Derived from date of entry to home. For Provider 2, only have month/year, so set to the 1<sup>st</sup> of the month. Data quality was improved across waves, since able to verify and correct anomalous dates (e.g. all set to the same date within a care home). Nevertheless, one case (n=1) omitted due to likely data entry error. |
| 4 | Textured food/diet | IDDSI 7 – regular<br>IDDSI 7 – easy to chew<br>IDDSI 6 – soft & bite sized<br>IDDSI 5 – minced & moist<br>IDDSI 4 – pureed<br>IDDSI 3 – liquidized | 687 | ≥71.3%<br>5.1%<br>9.5%<br>7.1%<br>6.0%<br>≤1% | 8.2% | 611 | ≥68.3<br>6.7%<br>8.8%<br>8.2%<br>7.0%<br>≤1% | 14.1% |  |
| 4 | Textured drink/fluid | IDDSI 0 – thin<br>IDDSI 1 – slightly thickened<br>IDDSI 2 – mildly<br>IDDSI 3 – moderately<br>IDDSI 4 – extremely | 672 | ≥89.9<br>4.5%<br>3.6%<br>≤1%<br>≤1% | 10.2% | 605 | 89.2%<br>4.8%<br>5.0%<br>≤1%<br>None | 14.9% |  |
| 4 | Cognitive impairment | Very severe<br>Severe<br>Moderately severe<br>Moderate<br>Mild<br>Borderline intact<br>Intact | 630 | 12.5%<br>14.4%<br>12.9%<br>17.9%<br>14.0%<br>9.4%<br>18.9% | 15.8% | 479 | 14.6%<br>17.3%<br>12.9%<br>18.6%<br>12.1%<br>7.7%<br>16.7% | 32.6% | Added to software for the MDS pilot.<br>The MDS CPS is calculated from five items: comatose, problem with short-term memory, cognitive skills for daily decision making, being understood by others, and eating ADL. Scored per Morris et al[41]. |
| 4 | Waterlow Score |  | 542 | 17.8, 7.3<br>(5 to 44) | 27.5% | 473 | 18.3, 7.3<br>(5 to 43) | 33.4% | Score of ≥10 indicates risk for pressure ulcer, with high risk ≥15 and very high risk ≥20. Full score range from 0 to 64. §§ |
| 4 | Braden Score |  | 345 | 16.6, 4.0<br>(7 to 23) | 53.9% | 277 | 16.4, 4.0<br>(8 to 23) | 61.0% | Full score range of 6 to 23, with higher scores indicating lower risk of pressure ulcers. Scores ≤10-12 indicate high risk and ≤9 very high risk. |
| 4 | Barthel Index |  | 582 | 41.5, 30.2<br>(0 to 100) | 22.2% | 288 | 34.9, 28.7<br>(0 to 100) | 59.5% | Added to software for the pilot. Score from lowest (0) to highest (2) level of functional independence for each item. These are summed, x5, to create a score from lowest (0) to highest (100) independence. |
| 4 | Delirium / I-AGED |  | 601 | 1.1, 1.8<br>(0 to 10) | 19.7% | 466 | 1.4, 2.1<br>(0 to 10) | 34.5% | Added to software for the pilot. Each of the ten items is scored no (0) or yes (1) and summed to create a scale from 0 to 10. Score of ≥4 indicates delirium[42]. |
| 5 | ASCOT Proxy- |  | 503 | .83, .19<br>(-.17 to 1) | 32.8% | 384 | .81, .19<br>(-.17 to 1) | 46.0% | Added to software for the pilot. Required some recoding to combine. |
|  | Resident |  |  |  |  |  |  |  | Applied preference weights for ASCOT SCT4 to generate index score from -.17 to 1.0 [43]. |
| 5 | ASCOT: anxiety and low mood |  | 503 | 4.0, 1.5<br>(0 to 6) | 32.8% | 403 | 3.9, 1.5<br>(0 to 6) | 43.3% | Added to software for the pilot. Required some recoding to combine. |
| 5 | ASCOT: Pain |  | 568 | 2.2, .8<br>(0 to 3) | 24.1% | 423 | 2.1, .9<br>(0 to 3) | 40.5% | Added to software for the pilot. Required some recoding to combine. |
| 5 | ICECAP-O |  | 583 | .73, .21<br>(0 to 1) | 22.1% | 300 | .71, .22<br>(0 to 1) | 57.8% | Added to software for the pilot. Required some recoding to combine. Provider 2: Data not provided for some residents. Score (0 to 1) calculated using UK index values [44]. |
| 5 | EQ-5D-5L Proxy 2 |  | 650 | .33, .35<br>(-.59 to 1) | 13.1% | 494 | .29, .34<br>(-.35 to 1) | 30.5% | Added to software for the pilot. Required some recoding to combine. Score calculated using the mapping function to convert to EQ-5D-3L and applied UK index values. The UK value set for the EQ-5D-5L is still being developed [45,46] |
| 5 | ASCS QoL | So good<br>Very good<br>Good<br>Alright<br>Bad<br>Very bad<br>So bad | 613 | 3.1%<br>26.3%<br>33.9%<br>28.9%<br>5.2%<br>1.5%<br>1.1% | 18.1% | 461 | 3.9%<br>25.8%<br>≥36.1%<br>27.3%<br>4.6%<br>1.3%<br>≤1% | 35.2% | Added to software for the pilot. Required some recoding to combine. |
| 6 | NEWS2/ RESTORE2 |  | 0 | N/A | 100% | 0 | N/A | 100% | Provider 1. Included in the software for data capture but no data entered by care homes (100% missing). Provider 2. Not available (100% missing). |
| 6 | MUST |  | 169 | .8, 1.2<br>(0 to 5) | 77.4% | 161 | 1.0, 1.3<br>(0 to 4) | 77.4% | MUST scored in last 6 months. A MUST score of 1 indicates medium risk and ≥2 indicated high risk of malnutrition. Not available for Provider 2 (100% missing). |

<sup>1</sup> **Wave 1:** n=748 (Provider 1 n=170; Provider 2 n=578)
<sup>2</sup> **Wave 2:** n=711 (Provider 1 n=168; Provider 2 n=543).
§ Included a flag to indicate cases with multiple entries (n=8) for Provider 2. As date of entry was unavailable, one entry was randomly selected.
§§ Included a flag to indicate cases with multiple entries for an individual (n=69) for Provider 2. As date of entry was not available, one entry was randomly selected.

Where data were already included in routine DCRs, some variables were more complete than others. CPR status was 99.6% complete. Care homes using Software Provider 2’s system did not routinely complete fields including marital status, first language, power of attorney and malnutrition universal screening tool (MUST), which contributed to high levels of missing data. For National Emergency Warning Score 2 (NEWS2) variables, no data were entered by care homes using either software.

The measures added to DCRs for the pilot were more consistently completed compared to those routinely recorded (Wave 1: <35% missing data). This is perhaps to be expected, since we required software providers to include these measures across participating homes, whereas homes could choose what routine data to record. We also devoted researcher time to explain the new variables and the rationale for their inclusion to care home staff.

In comparing Wave 1 and Wave 2, missing data increased by >8% for deprivation of liberty (7.9% to 21.2% for waves 1 and 2 respectively). For variables added to DCRs, missing data increased between 10% and 18% from Wave 1 to 2, except for the Barthel Index (increased by 36%) and ICECAP-O (increased by 37%). For Barthel, this was likely due to Provider 2 using a similar, but slightly different, version as their system default, which care homes reverted to using rather than the standardised version added for the study. Provider 2 did not return ICECAP-O data for five care homes at Wave 2.

Even with relatively high completion for QoL measures, there were issues with data quality in Wave 2. Provider 2 ‘carried over’ Wave 1 scores; therefore, care homes had to manually overwrite prepopulated scores. By contrast, Provider 1 required data entry of new scores for Wave 2. As a result, all but one care home using Provider 2 software had a maximum of two residents with any change in ASCOT-Proxy-Resident score between Wave 1 and 2, whereas only three residents had the same ASCOT Proxy-Resident score across waves for homes using Provider 1’s software.

### Accessed routinely collected health and social care datasets

We were able to retrieve and link data from PDS, SUS Admitted Patient Care, Outpatient and Emergency Care datasets, CQC care home data and supplement this with data from our online survey of care homes as planned. We were additionally able to collect data from the newly available national ambulance [47], adult social care client level [48], and community services (CSDS) [49] datasets. A care home residency table created by Arden & GEM Commissioning Support Unit [50] based on PDS data and estimated care home residency dates, and ONS Index of Multiple Deprivation data were also accessed.

Due to information governance constraints, a new data sharing agreement with NHSE was required, which was signed in October 2023. This delayed access to NHSE datasets and restricted the analysis possible in the remaining time. This also adversely impacted set up of data sharing with ICSs.

These datasets were accessed only for consented residents and not for all care home residents in the ICSs as originally planned [11]. In addition to IG challenges, this was primarily because the underlying flow of data previously used to identify care home residents had been replaced, resulting in the complex algorithm [45] for care home identification needing to be redeveloped and validated by NHSE.

We were unable to access GP records because we couldn’t establish data sharing agreements for two of the ICSs in time for the study. In the remaining ICS we were able to secure some data sharing agreements with GP practices by working through a Commissioning Support Unit (CSU), a regional body providing data support to NHS organisations. However patient data are held by individual GP practices, and we had to liaise with multiple Data Protection Officers within the same ICS. Ultimately, the number of resident records available from GP practices that signed agreements in time was too low to ensure residents could not be re-identified, and therefore it was not possible to proceed to extraction under General Data Protection Regulations (GDPR). A list of data items we would have accessed from one ICS where we established data sharing agreements, had we been able, is available in Appendix 7.

The inability to collect GP data was a major contributor to the differences between the aspirational and final prototype MDS, summarised in Appendix 8. Other contributors were poor feasibility of extraction from DCRs and high levels of missing data for some items in routine datasets, rendering reliable counts of activity linked to particular conditions or events impossible.

### Creating derived variables in prototype MDS

Due to the absence of GP data, comorbidities were derived from SUS data, using a 3- year lookback period from the index date. We couldn’t derive these for 144 residents (20%) who didn’t have a hospital admission in that period. Activity summaries were reported for the year leading up to the index date, independent of whether residents joined their current care home within this time period. On average, residents in Wave 1 had been living in the current care home for 28.7 months, with 29% having moved in within the year leading up to their index date.

### Hierarchy process

Table 3 presents the variables included in the hierarchy. For universally defined variables, there were high levels of consistency where recorded. Levels of completeness varied widely – from 1% missing for sex in CSDS to 80% missing for ethnicity in the care home record. Overall, the process of using information from several sources to populate the final variable included in the MDS greatly reduced the level of missing data (missingness <=4% across variables).

**Table 3.**
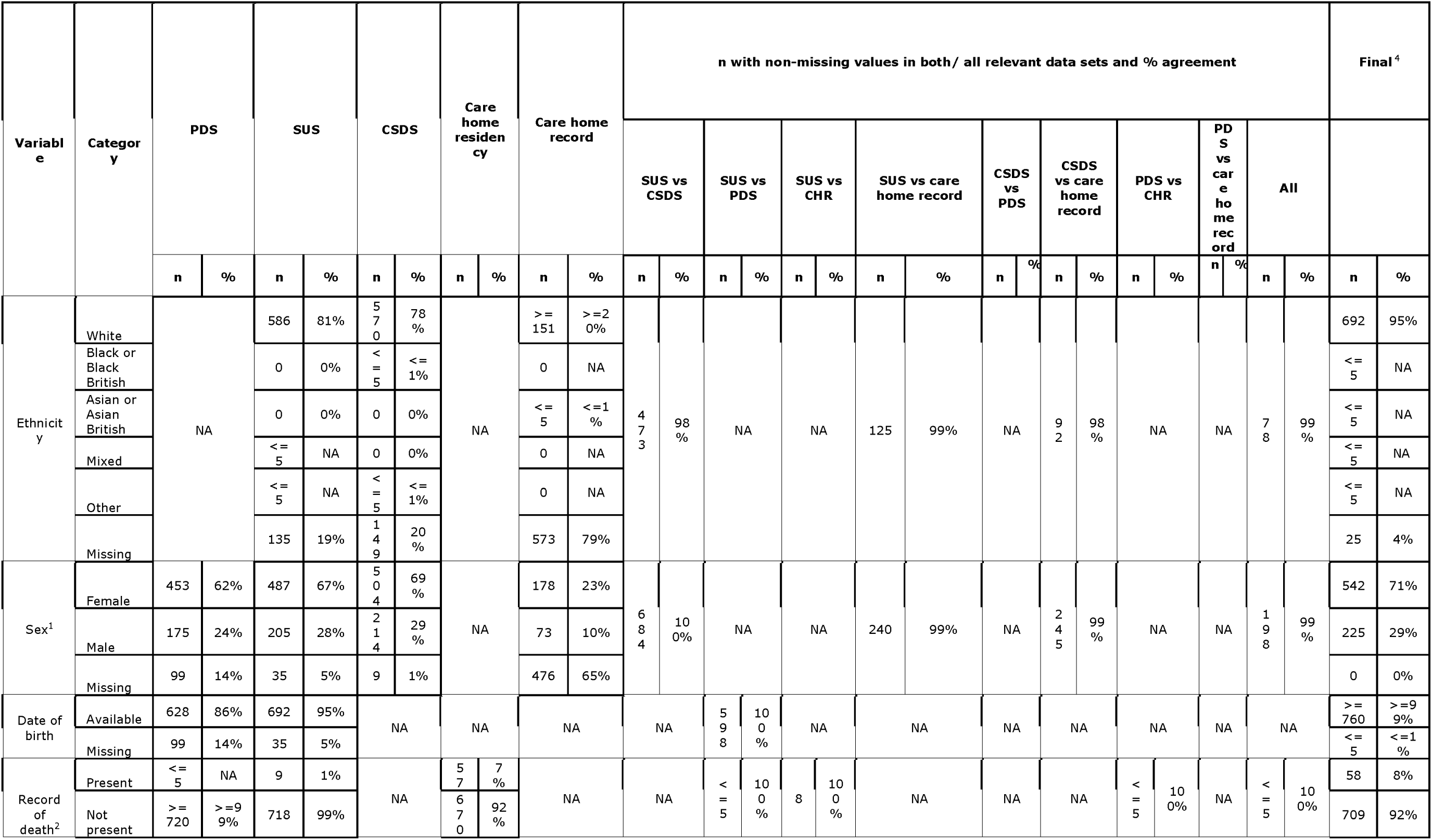
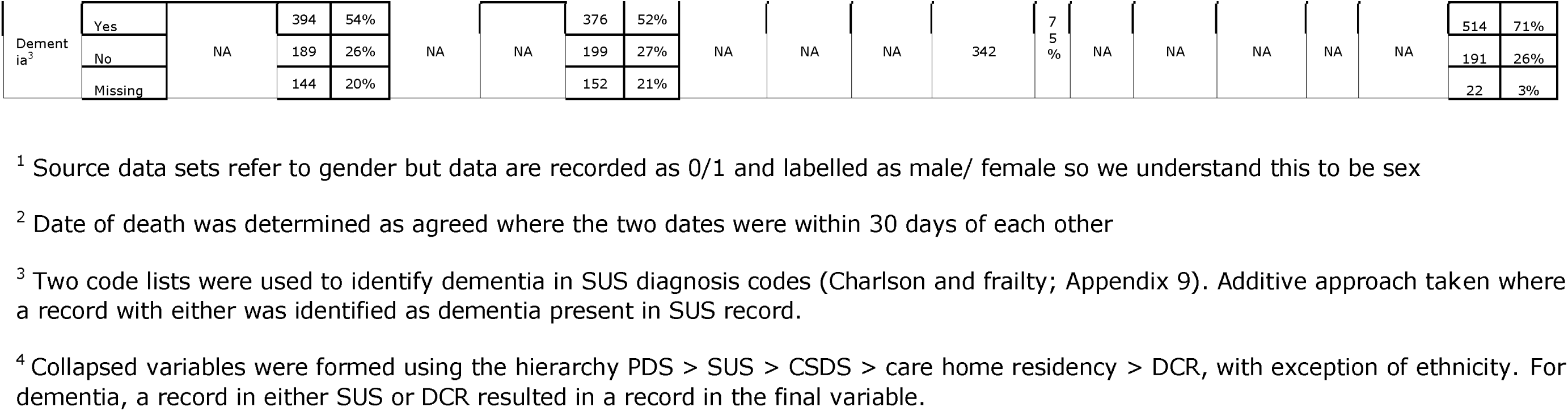
Comparison of variables across data sources to determine hierarchy.

### Final prototype MDS

Key variables from the final prototype MDS are summarised in Table 4. Appendix 10 shows the full version, which includes two approaches to healthcare utilisation – mean activity across all residents, and proportion of residents with at least one event.. Appendix 11 contains worked examples, based upon our work with stakeholders, of how data from the MDS could be used to help understand Emergency Department and Ambulance contacts.

**Table 4.**
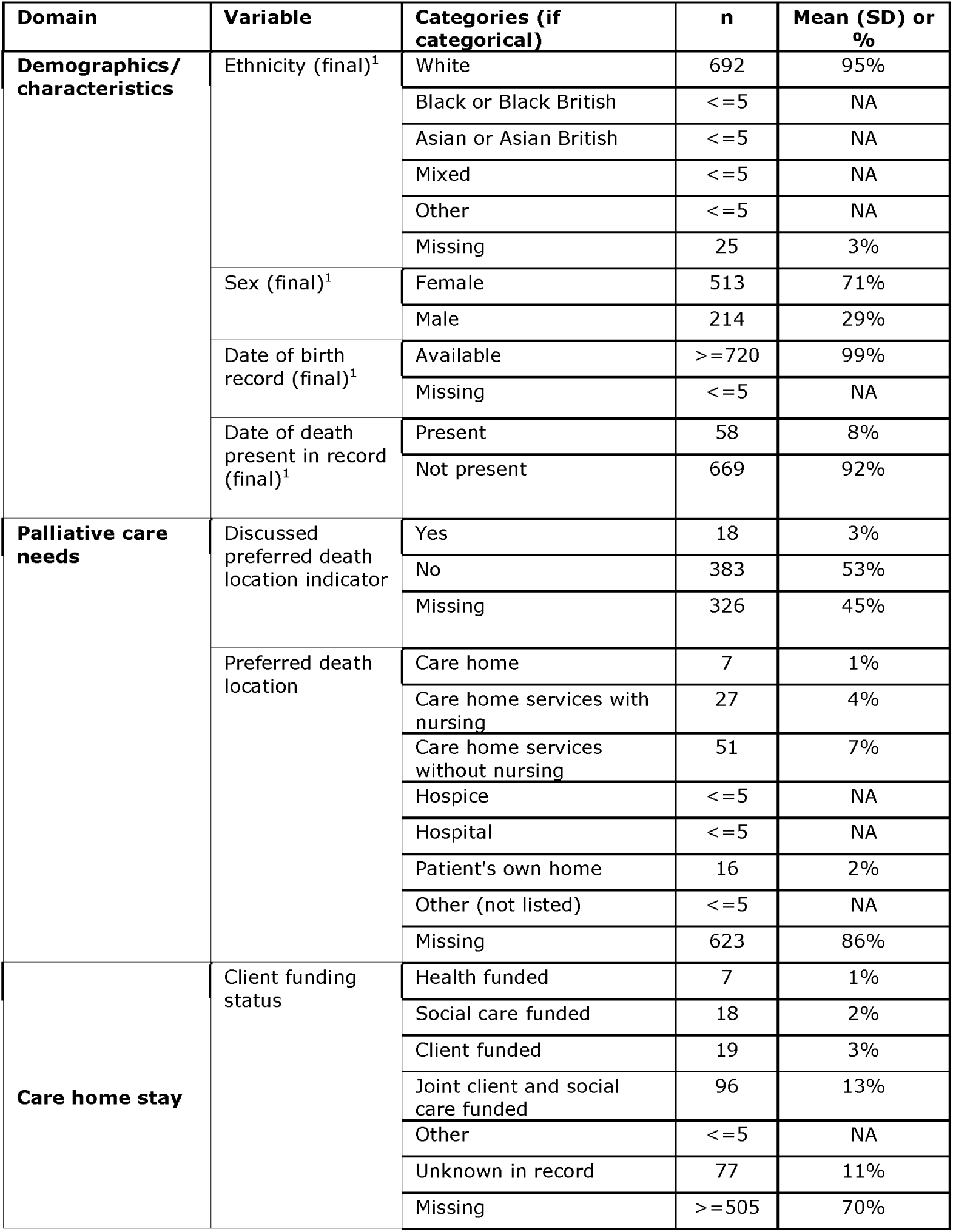
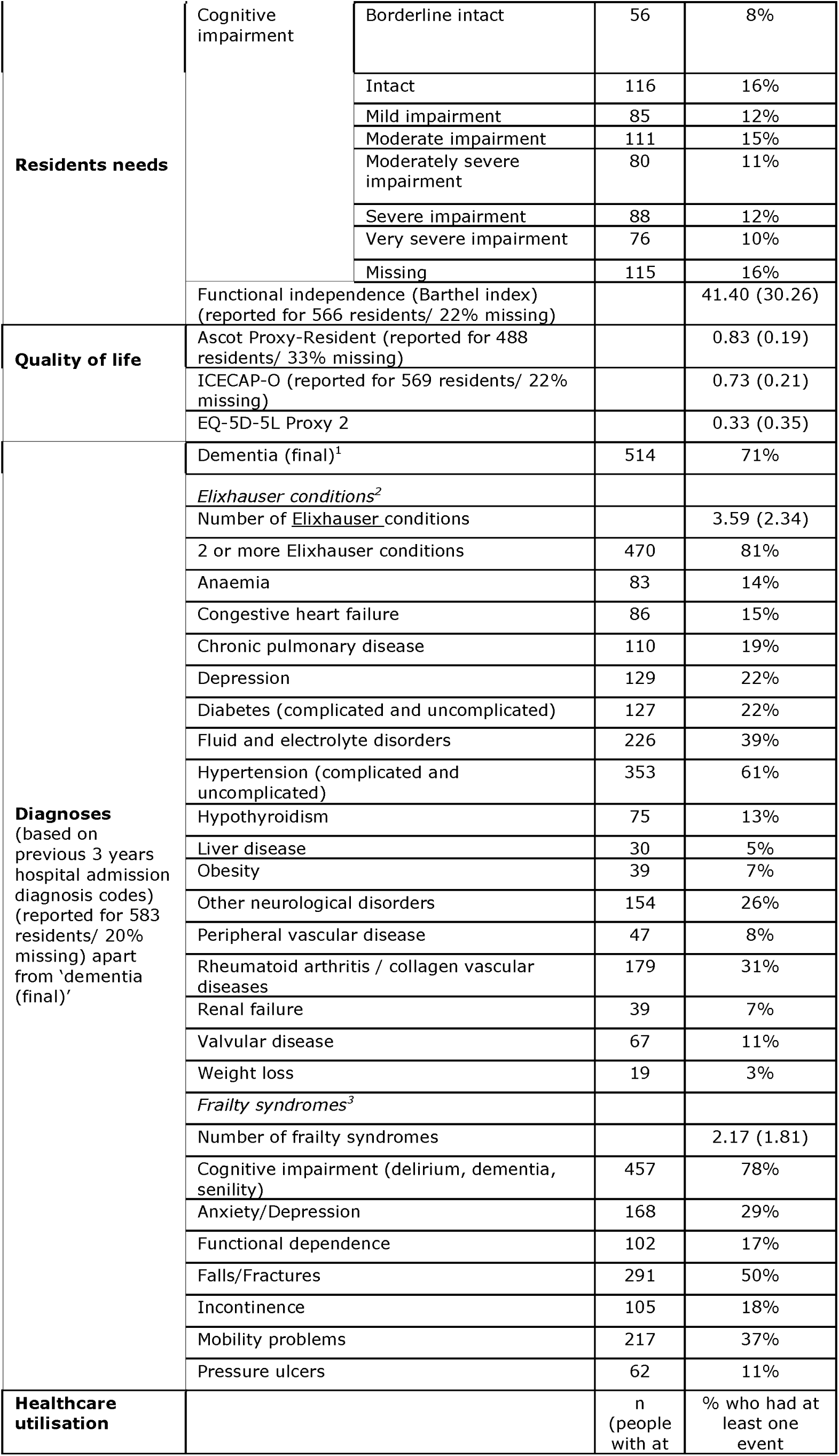
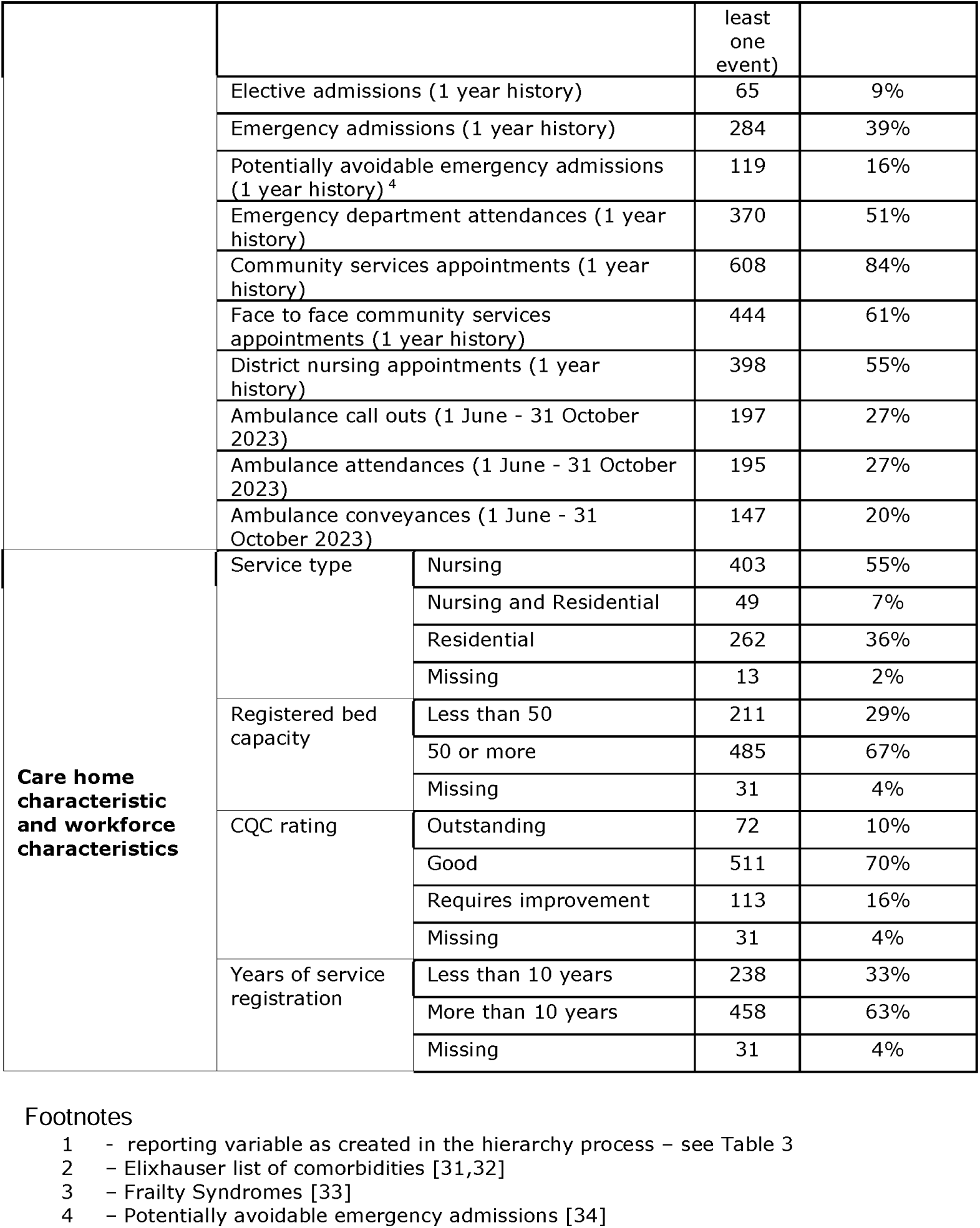
Selected variables from final prototype MDS. Numbers are reported for 727 residents unless otherwise specified.

## Discussion

In the face of substantial challenges, many of which were not unique to this study[51,52], we accessed information from care home DCRs and safely linked data from multiple sources and data owners to create a viable prototype MDS for English care homes. Our prototype MDS was cross-sectional. Real-world deployment would be longitudinal, with data extracted at regular intervals, balancing the requirements of those funding, planning and delivering services against burden of data completion and collation.

We set out to collate routine administrative health and social care data for all care home residents in participating ICSs, with linkage to DCRs taking place only for those giving consent. This should have been technically feasible using methods outlined in this paper alongside a published algorithm to identify care home residents in routine data [53]. However, the algorithm was under redevelopment at the time of our pilot and couldn’t be validated in time to be incorporated in our data flow. Our final prototype MDS was therefore limited only to residents providing consent to linkage. This may have introduced systematic bias and data presented here should not be seen as representative of the wider UK care home population. For example, our data on healthcare resource use should be interpreted with caution – we do not know how health status influenced ability to provide consent.

Our data on health status, meanwhile, are limited by lack of access to GP records. This is reflected in lower reported prevalence of common conditions, such as dementia, than in previously published studies, although the prevalence based upon MDS CPS corresponds better to the prevalence cited elsewhere [1,54]. Long-term conditions such as incontinence and hearing loss, central to understanding healthcare needs in care home residents, are under-recorded in secondary care records [54]. If the MDS presented here is to be of use in practice, incorporating GP data is essential. The challenges encountered accessing GP data related to information governance and our role as researchers external to the ICS, coupled with time constraints. It was not due to resistance to the principle of data linkage. GP practices work as independent contractors commissioned by the NHS, each practice acts as data controller for their own patients’ data and there is as of yet no national GP dataset.

Our design repurposed routinely collected care home data to minimise care staff burden and focus on capturing what was important to staff and residents. Where data were central to routine care delivery – such as CPR or Deprivation of Liberty status – they were largely complete. Variables that were incomplete were either: regarded as superfluous because care staff know these for their residents (e.g. ethnicity or marital status); captured in free text and difficult to analyse; or difficult to record in a dependent population, (e.g. weight). Variables added via external mandate (e.g. NEWS2, included at the request of healthcare providers) [55], were not completed. For variables added to DCRs by our research team for the pilot, we saw initial high completion rates fall during the second wave of data collection. This was multifactorial, with competition for staff time, staff attrition, and implementation issues including a duplicate Barthel index in some care homes’ software, all contributing. These findings align with previous research on the importance of understanding the context of data collection when working with and interpreting data from social care [4,56].

An “off the peg” internationally validated MDS would not necessarily be a viable alternative for the UK. It superimposes a new system of data capture onto care homes, not linked to health and social care data held elsewhere and favours health data over quality of life and social care data. The issues we addressed around GDPR, the labour intensive and manual nature of linkage between care home and NHS data, the complex hierarchy of statutory databases into which an MDS has to interdigitate, and the need to train and invest in care home staff over time, would be the same. Previous attempts to use such MDSs in UK research studies found low completion rates and crucially, a higher burden associated with staff completing them on top of existing data requirements [57,58]. The work of implementation for uptake and sustained use is as significant and arguably more resource intensive than we found for our prototype dataset[59].

We faced issues with standardising approaches to data collection across two software providers and forty-five care homes. Plans underway by the Department of Health and Social Care to develop a Minimum Operating Data standard (MODS) [10] might facilitate some standardisation going forward. However, this MODS has been designed without the comprehensive evidence review and stakeholder consultation conducted for our pilot, and it contains a fraction of the variables included in our prototype MDS. It is likely to be at best an adjunct to a more comprehensive solution and will likely require iteration as implementation challenges, of the sort described here, unfold.

Our prototype MDS focussed on healthcare variables. This reflects, in part, the prominence given to these by all contributors, including care home staff and public representatives, during stakeholder work [16–18]. It also reflects the fact that routine healthcare data are often collected in a way that enables systematic collation and linkage. We found some data in DCRs stored as free text – an approach that provides nuanced and personalised records but hampers collation and analysis at meso- and macro-levels. The incorporation of social care related QoL and wellbeing, in the form the ASCOT-Proxy-Resident and ICECAP-O measures, goes some way to bridge this gap by providing person-centred data focussed around what matters to residents and relatives, collected in a standardised way. QoL data have been highlighted as essential for understanding quality in the sector[60]. For now, there is a trade-off between data collatable in an MDS and data held in free-text. This may, though, be addressed by advances in machine-based analysis of free-text in the future.

We presented in an appendix how the MDS could facilitate understanding of care home residents’ use of ambulance services and hospital emergency departments. Our stakeholder work revealed other areas where an MDS could generate insights, including reasons for hospital admissions to inform local service provision or training needs, and understanding pathways and access to services for residents with, for example, diabetes or mental health needs. Whilst this stakeholder wish list demonstrates the potential of an MDS to better understand resident needs, it also raises the challenge frequently reported in the care home literature, of care home staff and providers feeling that they are at the mercy of external forces beyond their control [4,12,58,61]. The evidence on what enables NHS services working with care homes to achieve improved outcomes consistently points to systems and practices that initiate and sustain quality of working relationships between health and social care staff and their organisations [58,62–66]. The powerful insights deliverable through an MDS come with attendant responsibilities. Ensuring that data are used in a way that foster trust between different stakeholder groups is an implementation imperative.

In conclusion, we have developed and demonstrated an MDS based on data-linkage for English care homes. We have identified issues around data quality, information governance, plurality of data and the need for implementation approaches that facilitate data completion, that are essential to implementation of any MDS in English care homes. We have also demonstrated the value of combining data sources to provide richer data and crucially reduce external requests for information from care homes. It is essential that this work moves forward to ensure that we can take data-informed approaches to care delivery, service design, commissioning and policy for the care home sector.

## Ethics approval

The study has received ethical approval from the London Queen’s Square Research Ethics Committee (22/LO/0250).

## Data availability

Pseudonymised data will be available on request from the corresponding author following a 24 month embargo from the date of publication.

## Acknowledgements

With thanks to Melissa Co, Sarah Opie-Martin and Tom Prendergast for their help with data cleaning, analysis and quality assurance of the analysis code, to James Lockyer for helpful discussions about routinely collected health and social care datasets, and to Sarah Hardy for her help with final write-up and proof-reading.

## APPENDIX 1 Core Tenets of an MDS for long-term care homes, reproduced from Burton et al[20]

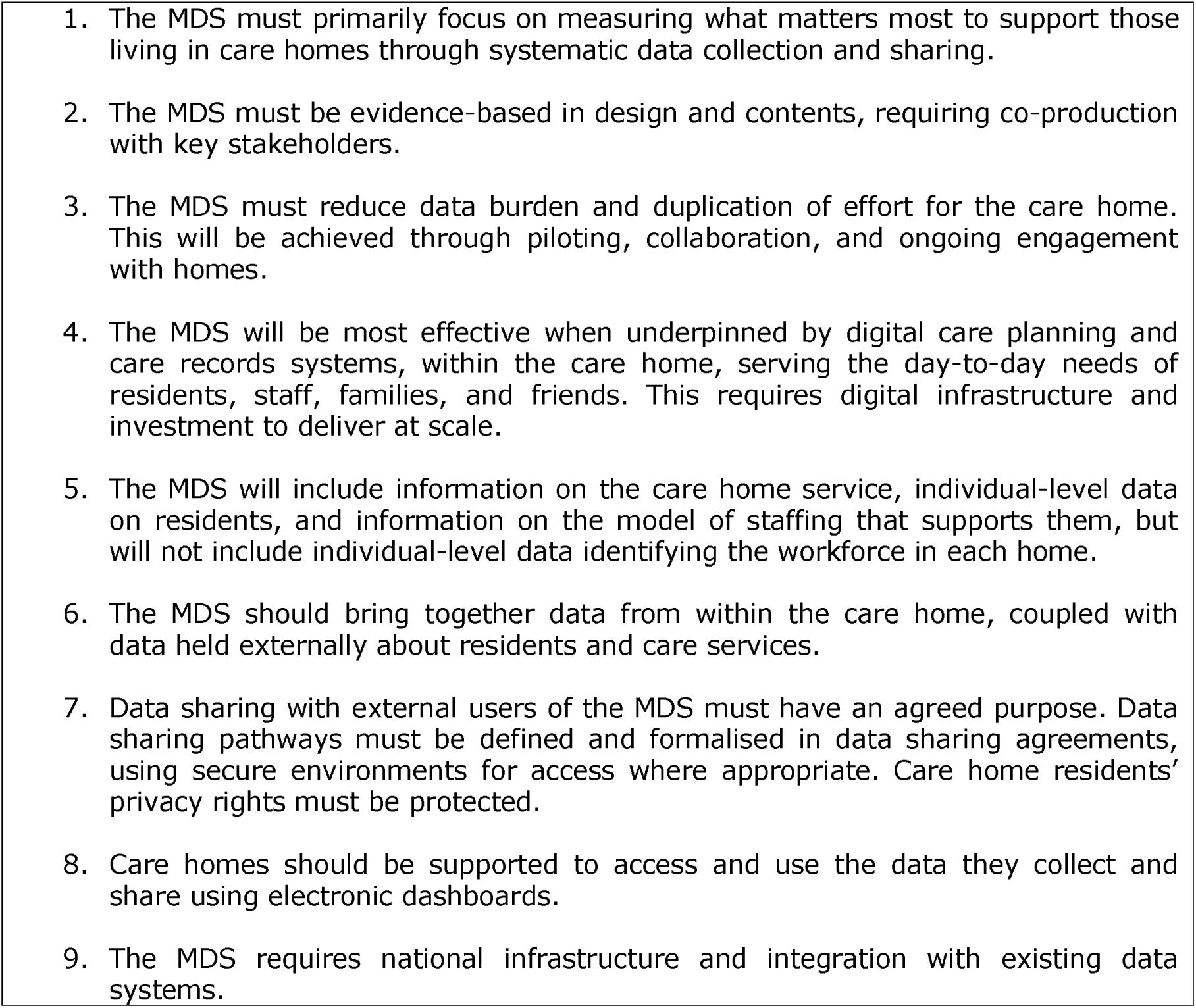

## APPENDIX 2 the Aspirational Minimum Data Set with Proposed Dataset as published in the initial study protocol [11]

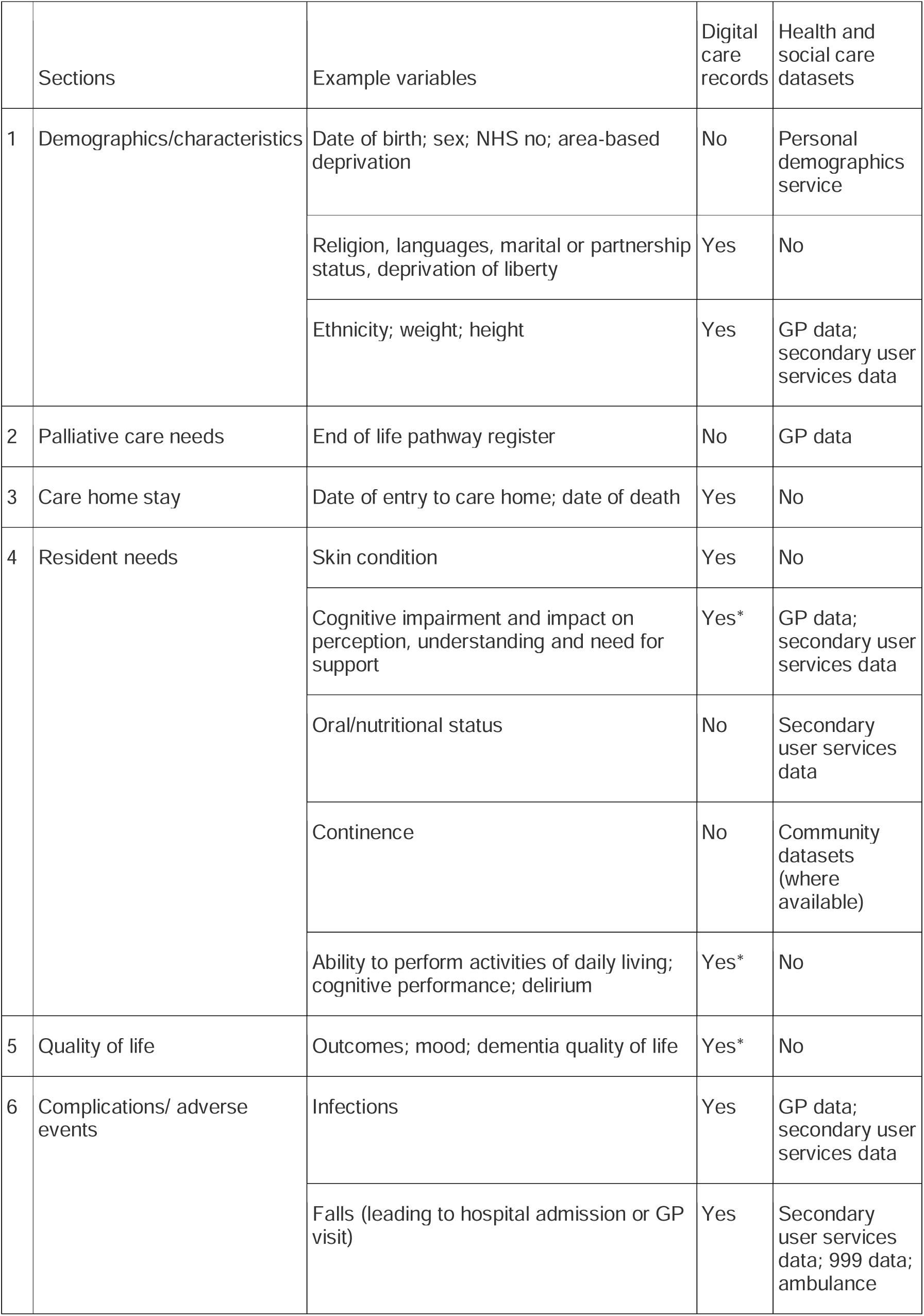
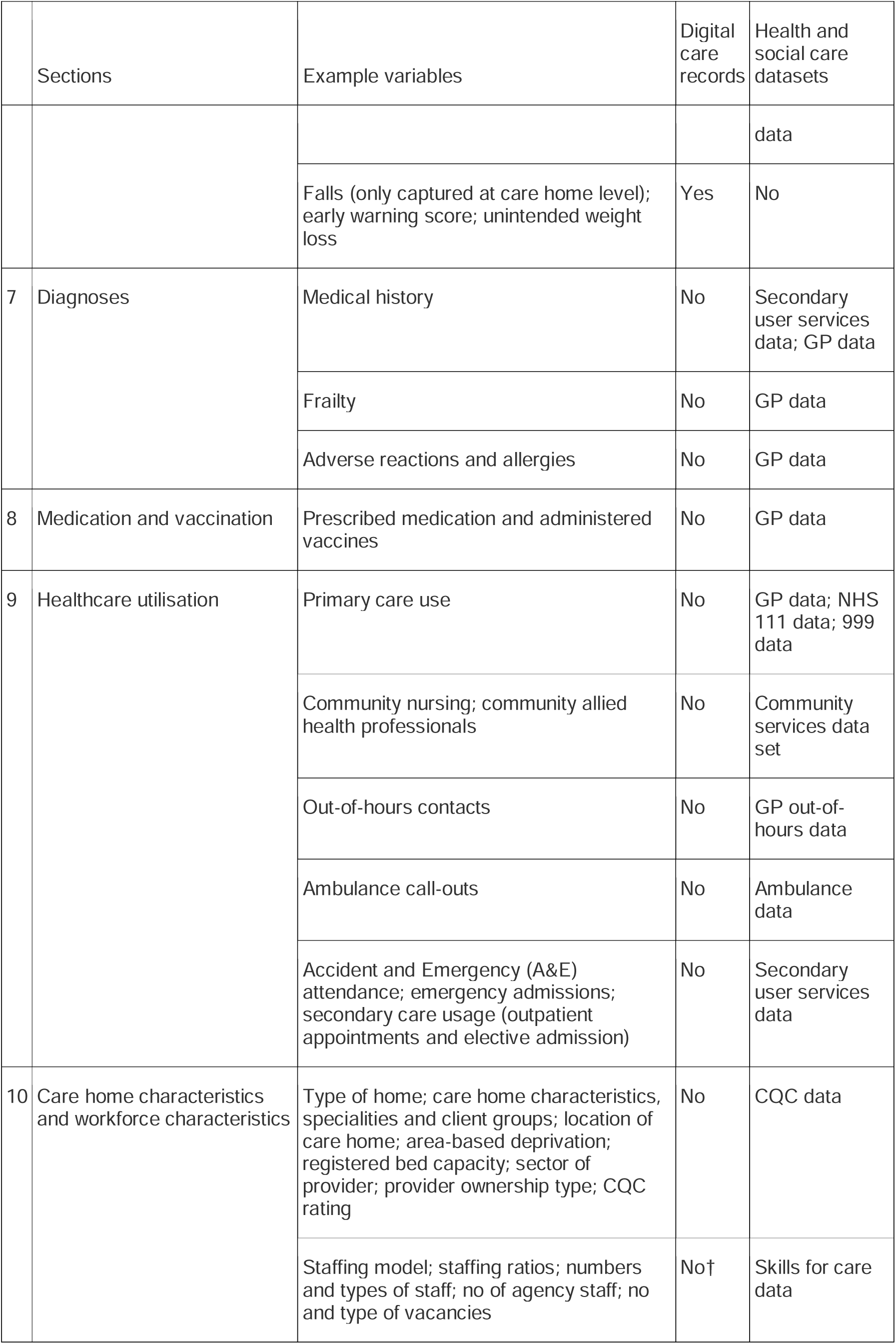
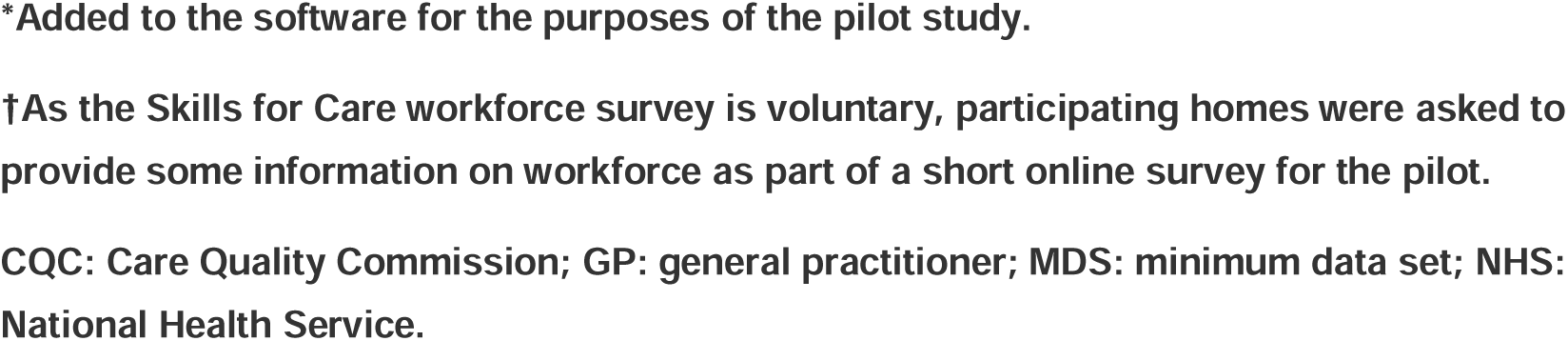

## APPENDIX 3 Data Flow Diagram

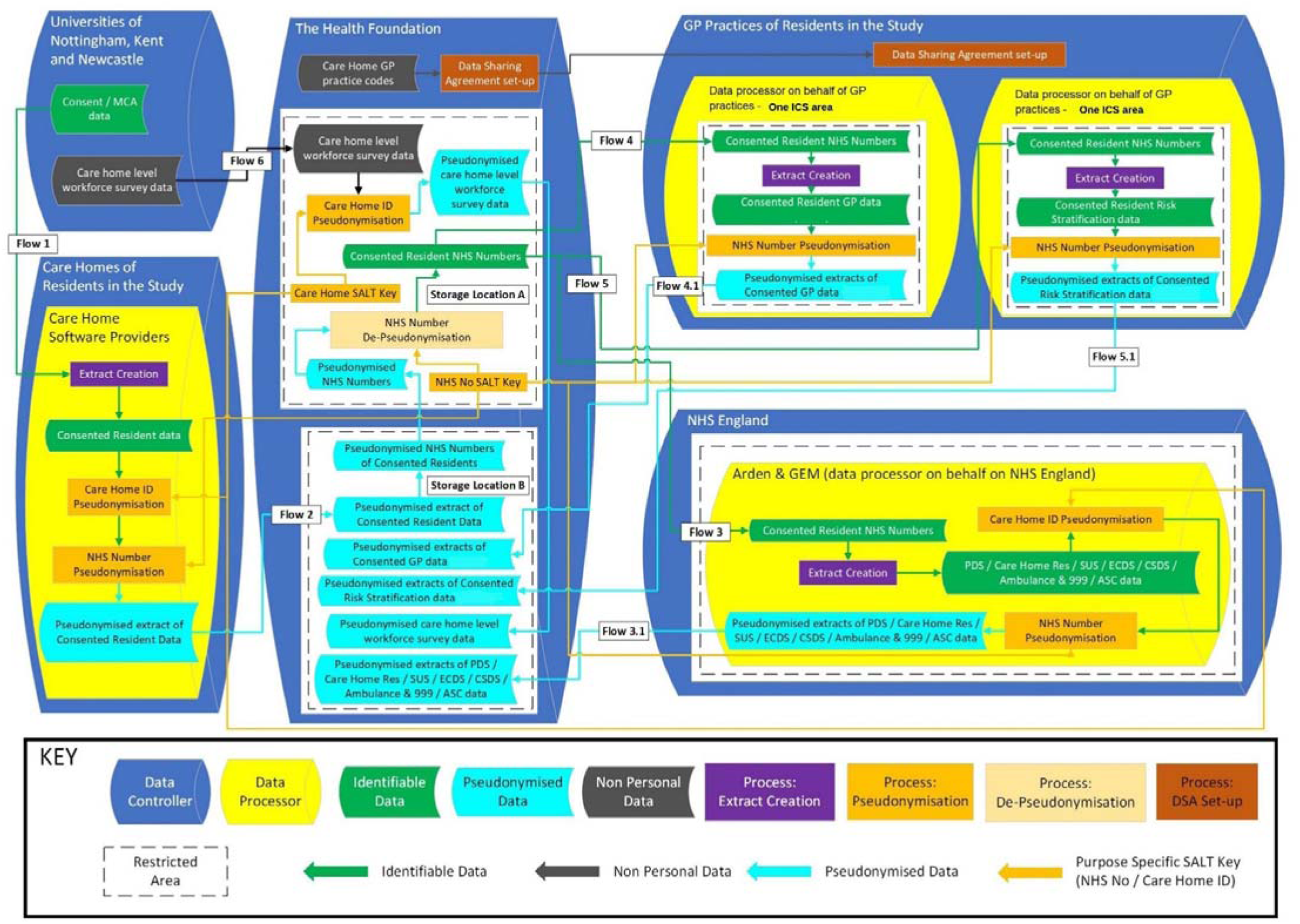

## APPENDIX 4 Data Sharing Summary Diagram

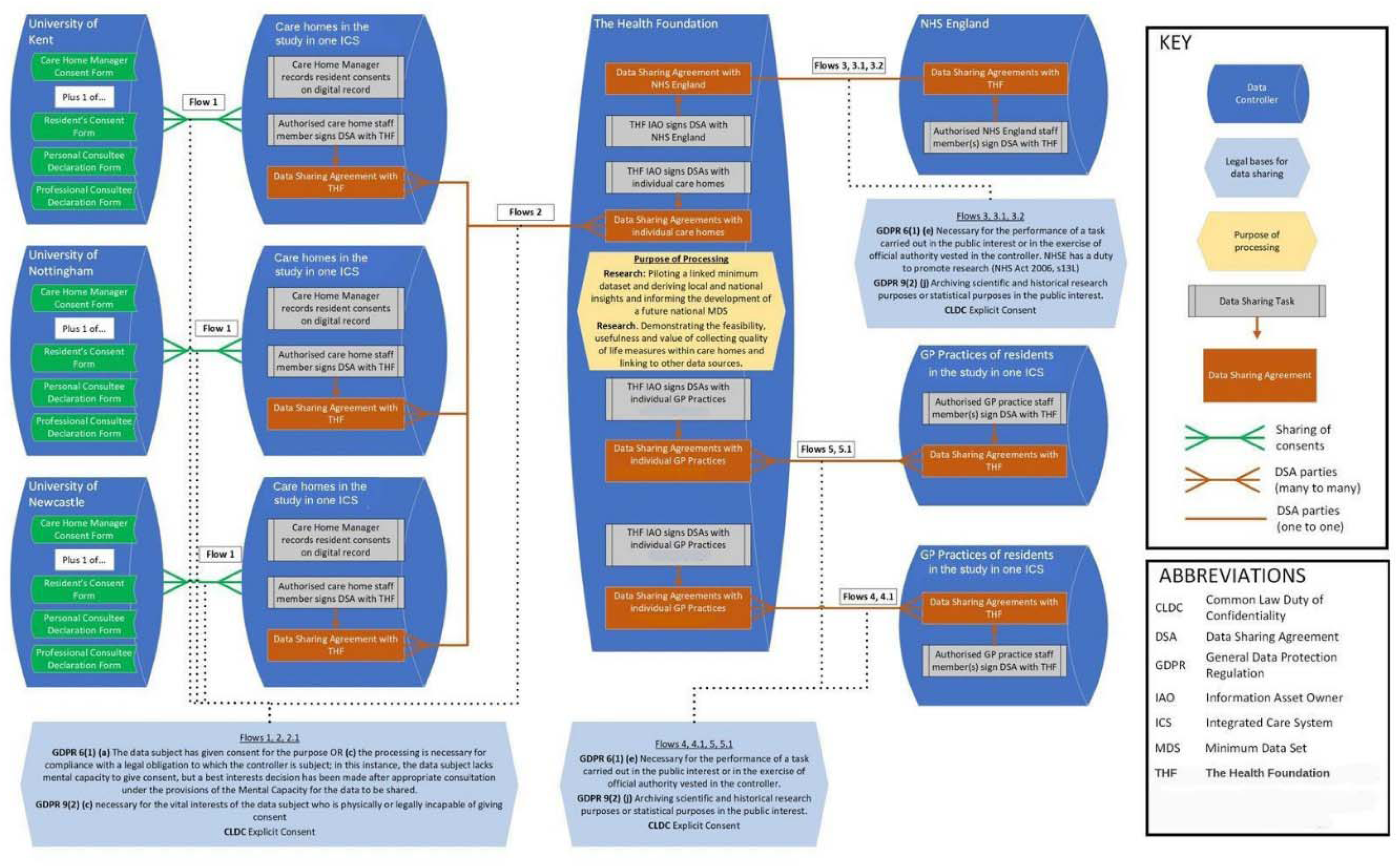

## APPENDIX 5 Data specifications

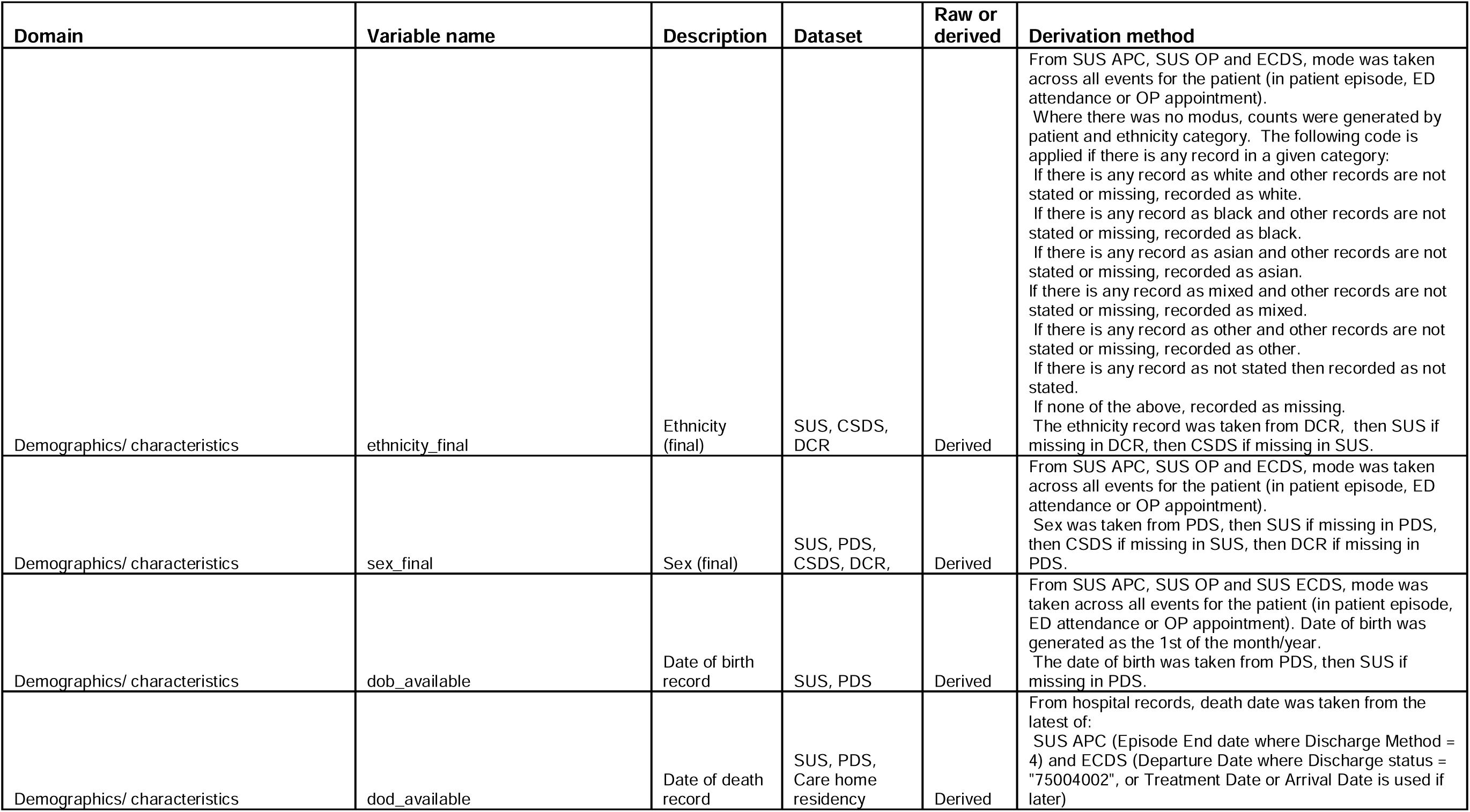
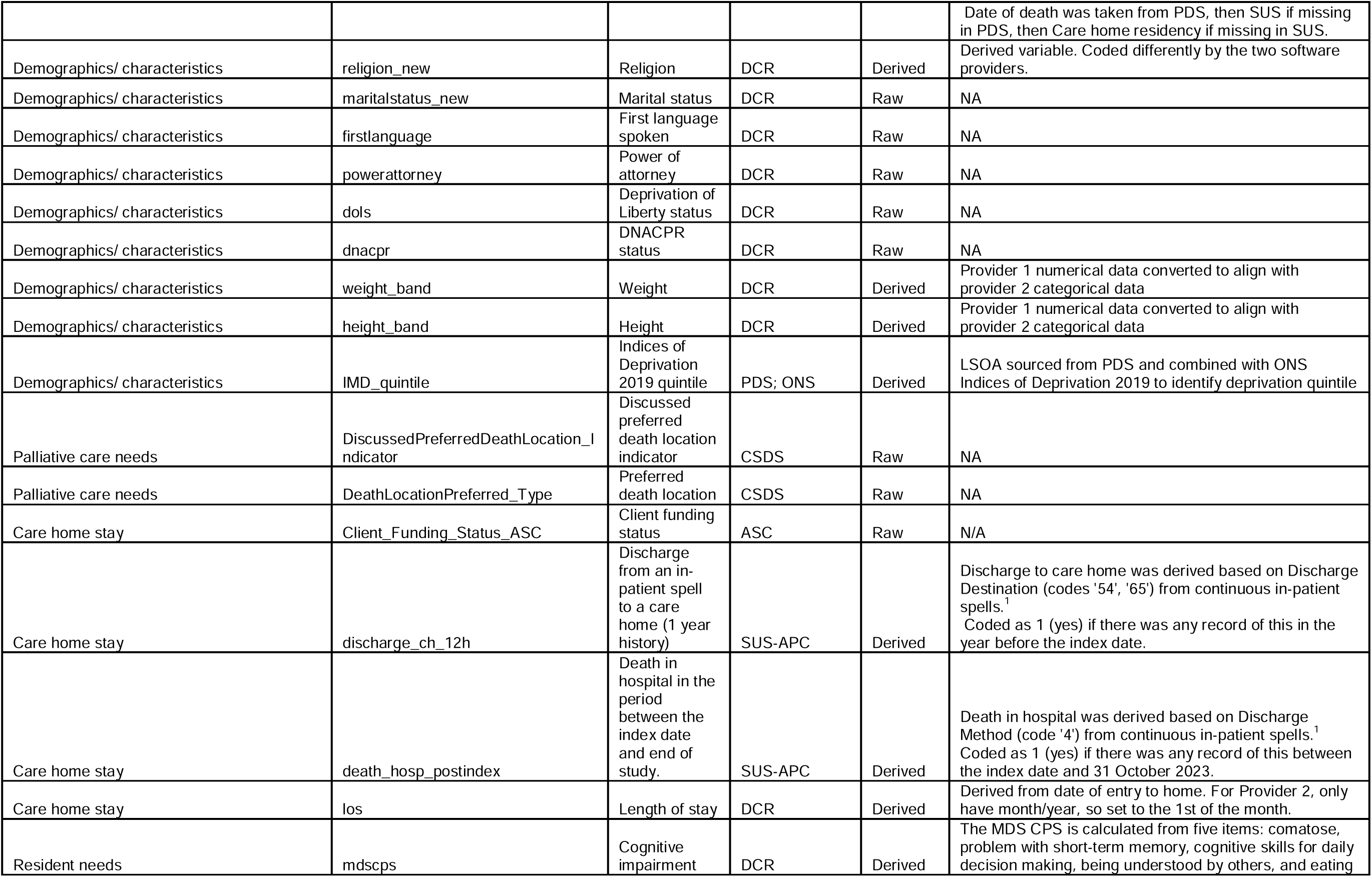
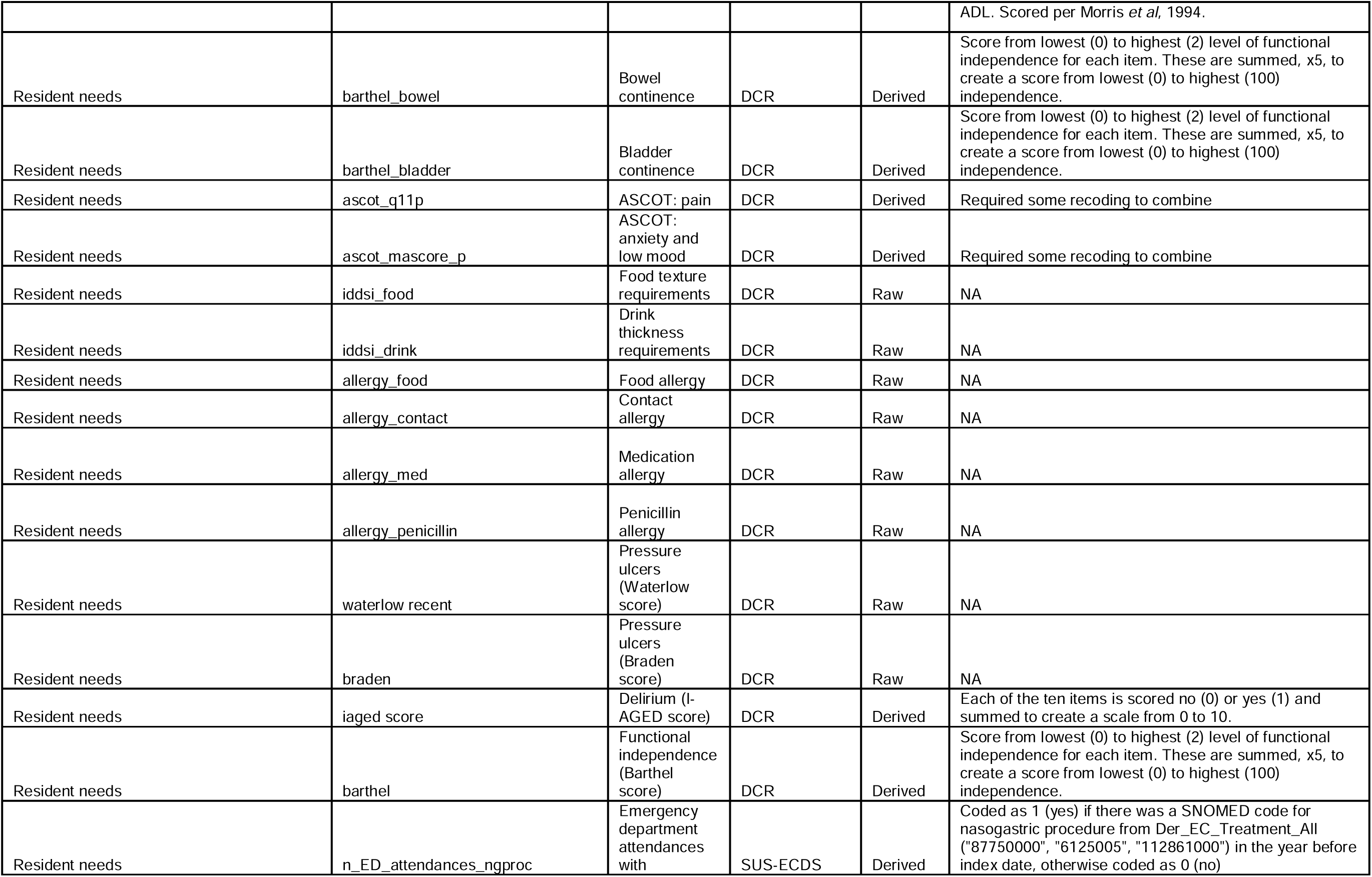
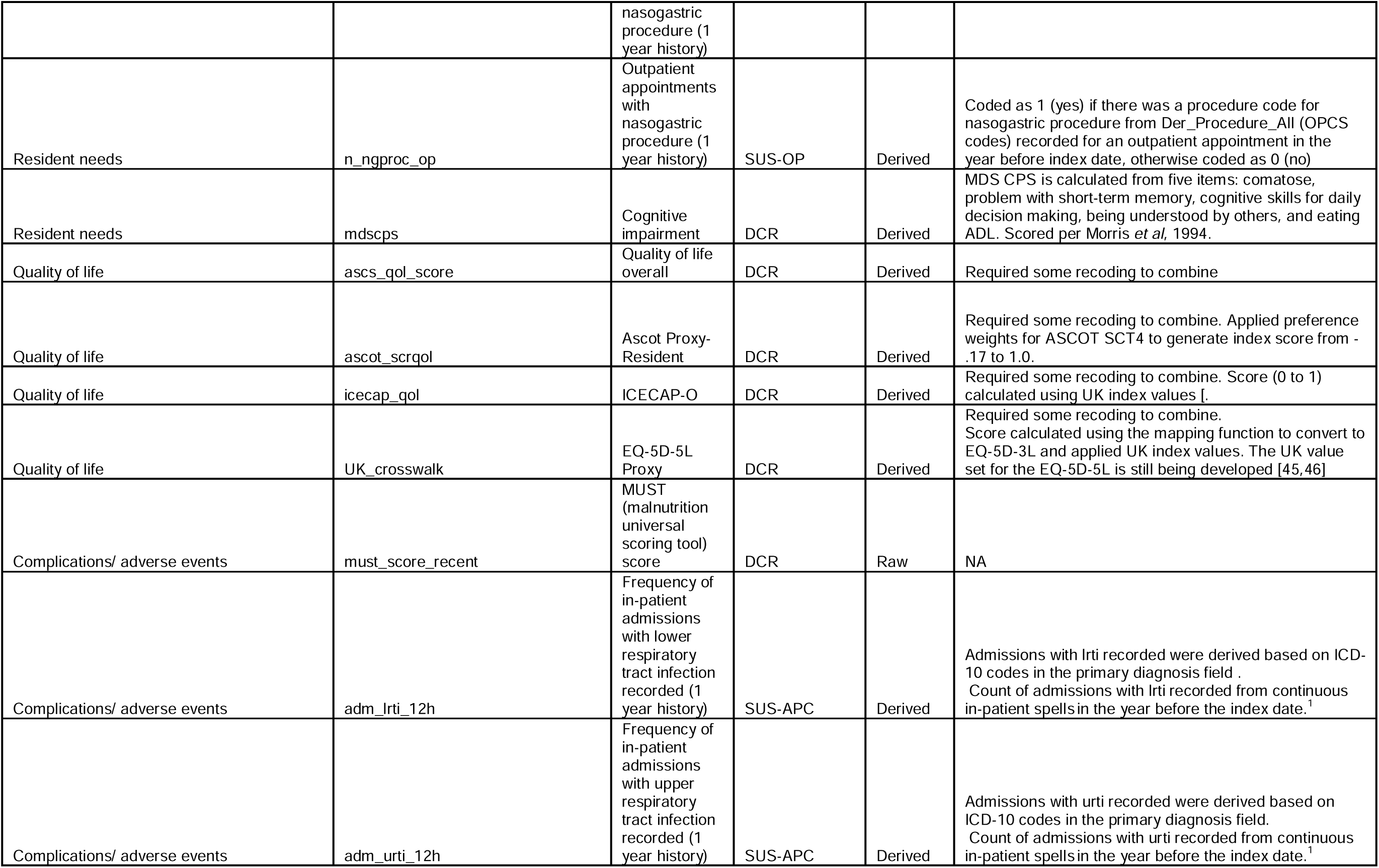
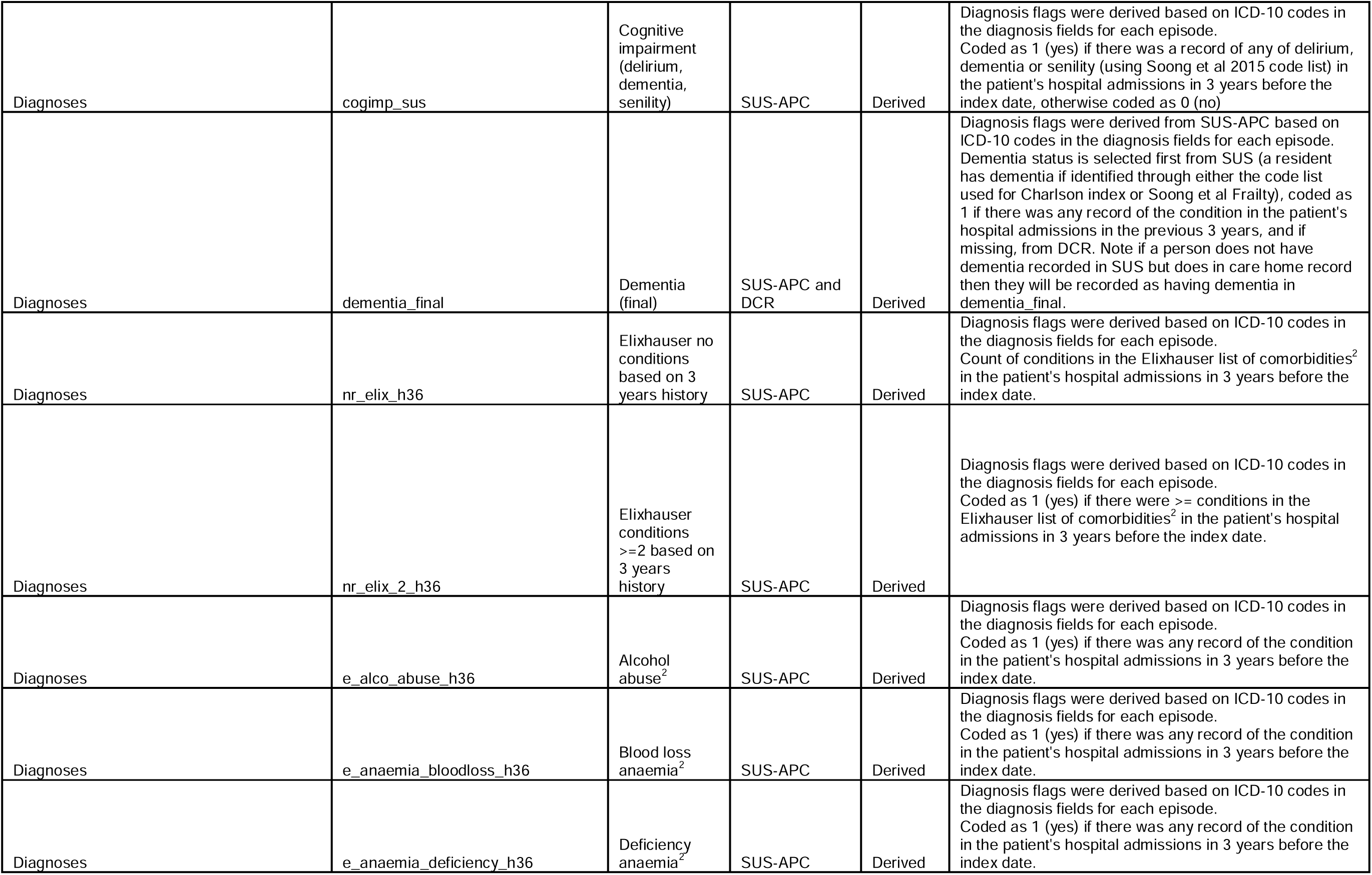
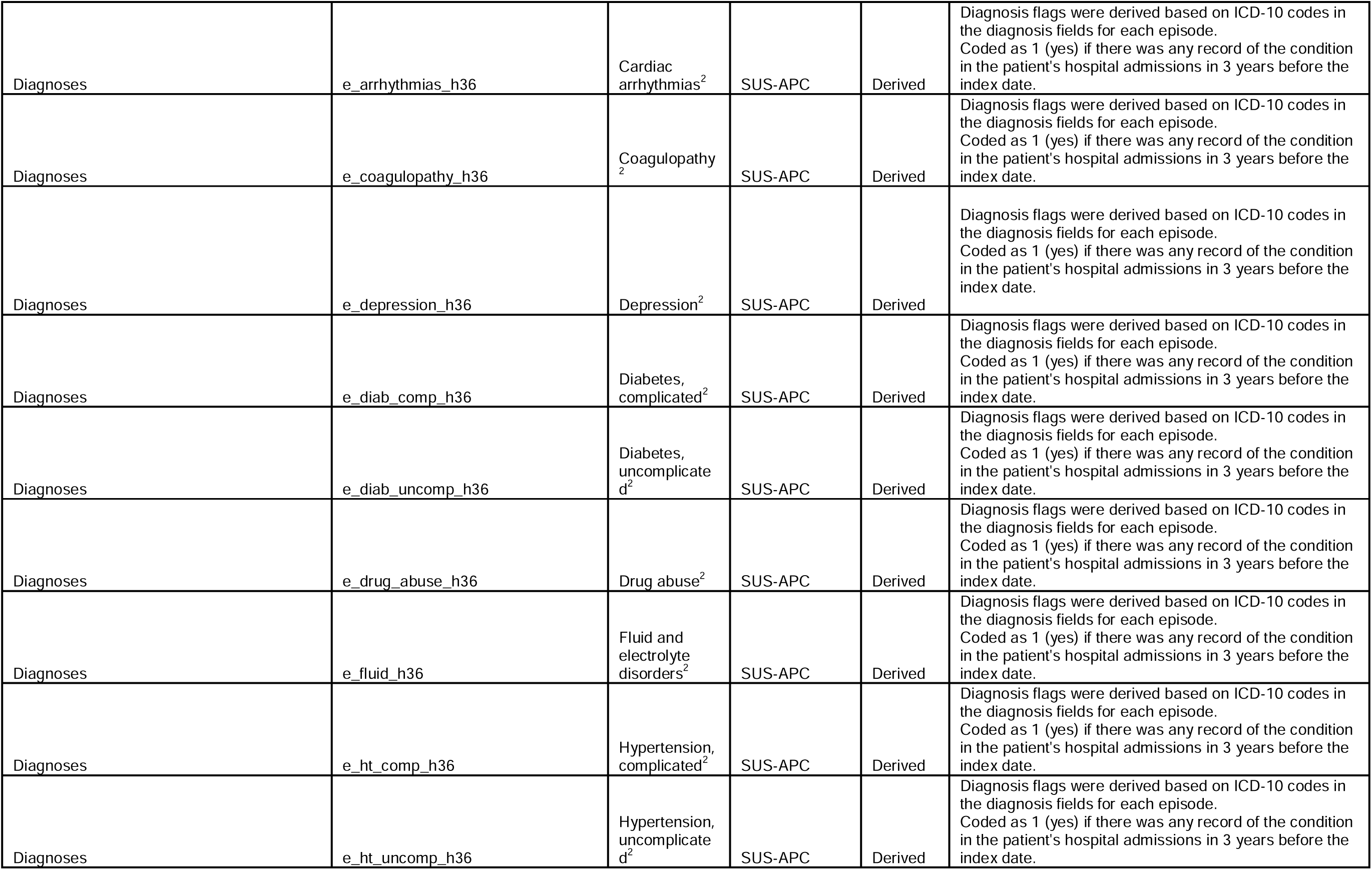
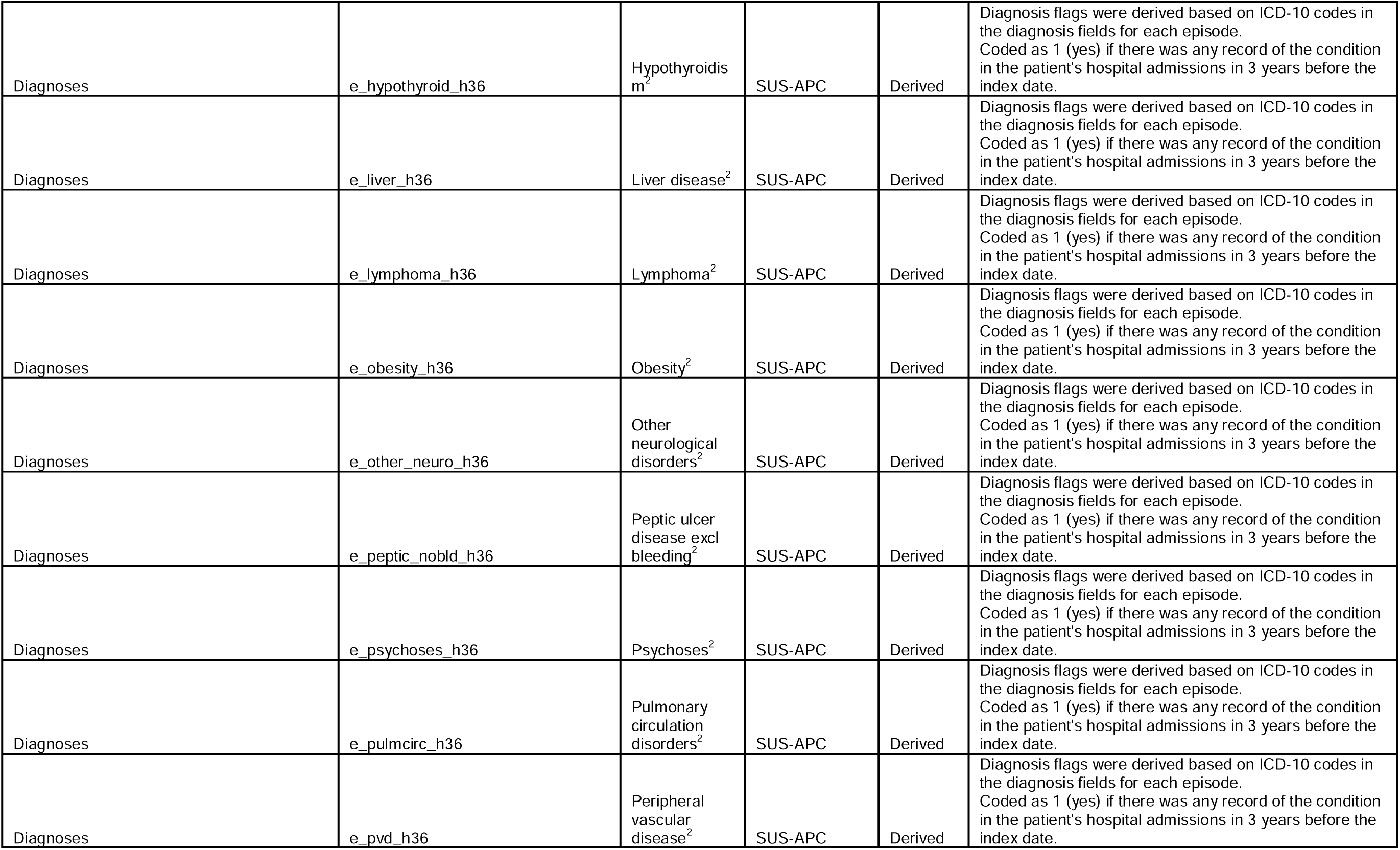
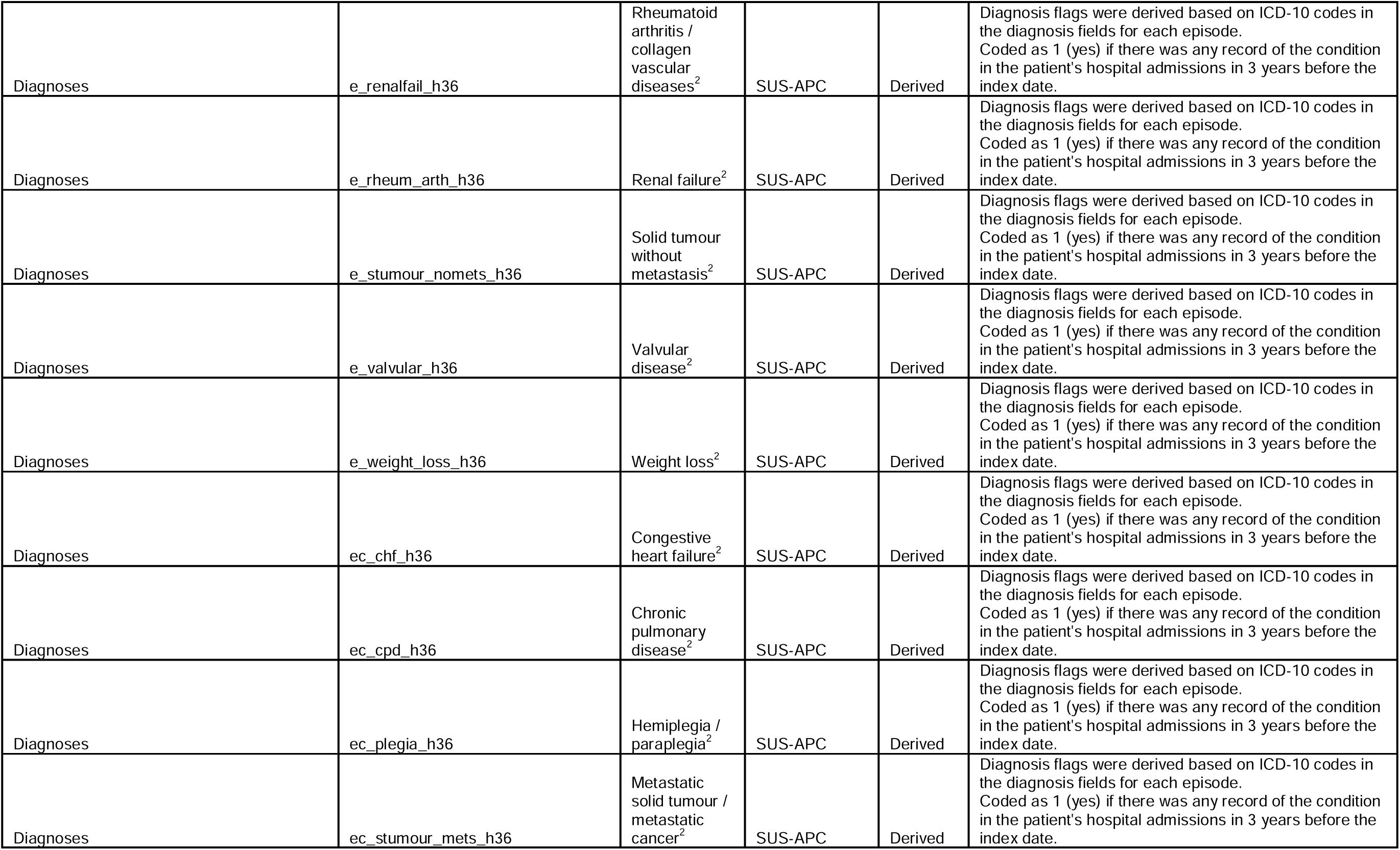
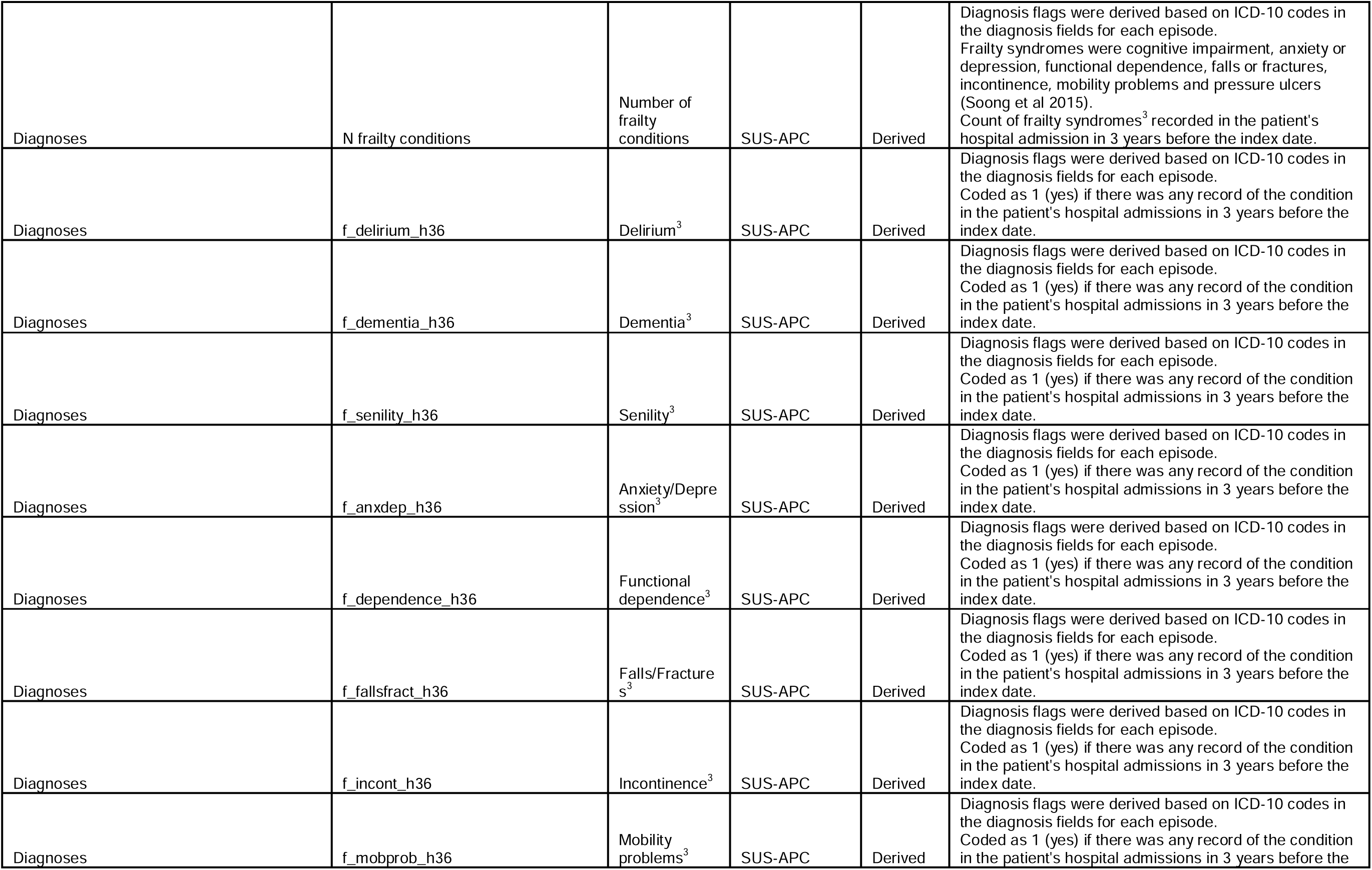
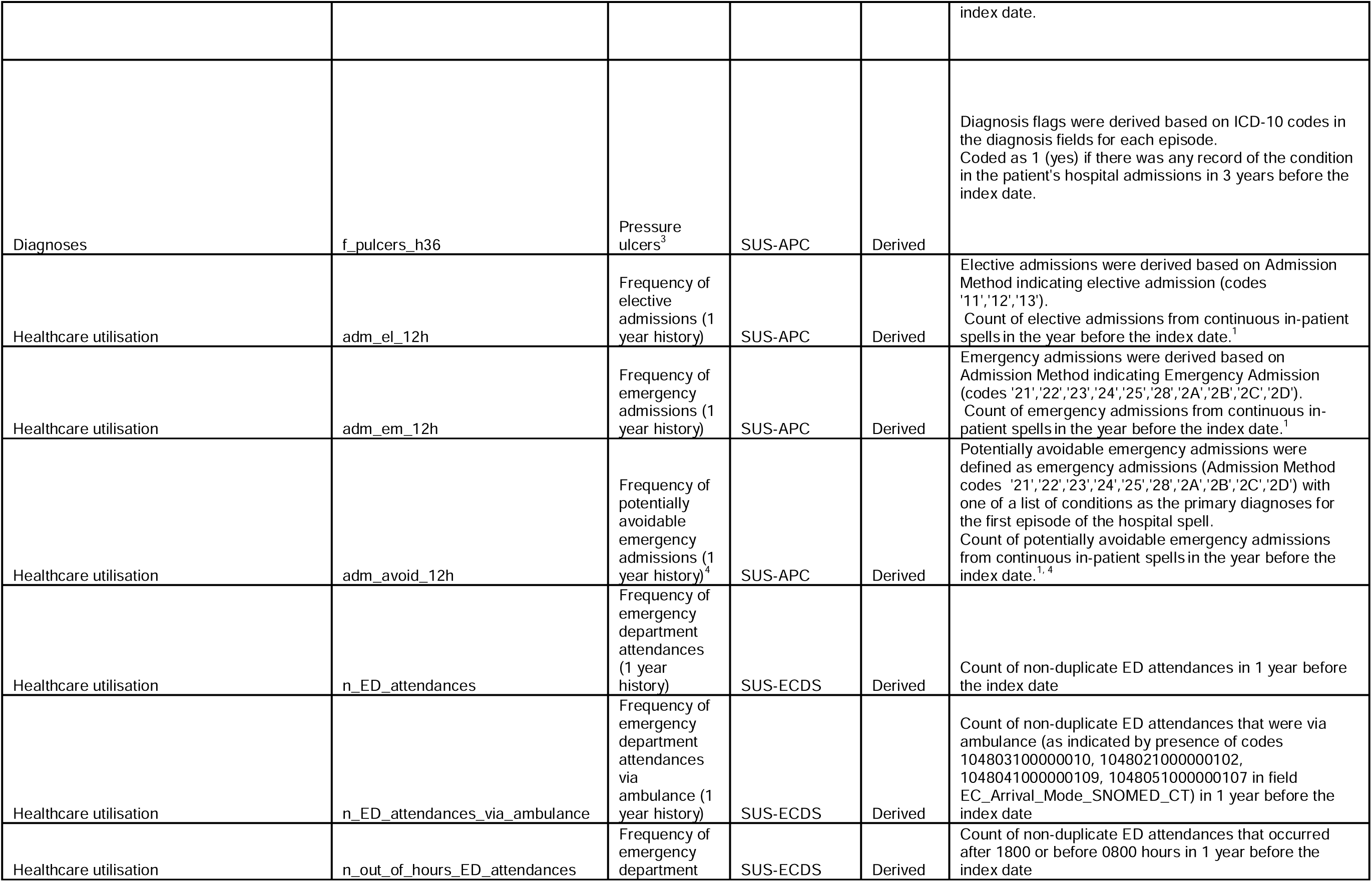
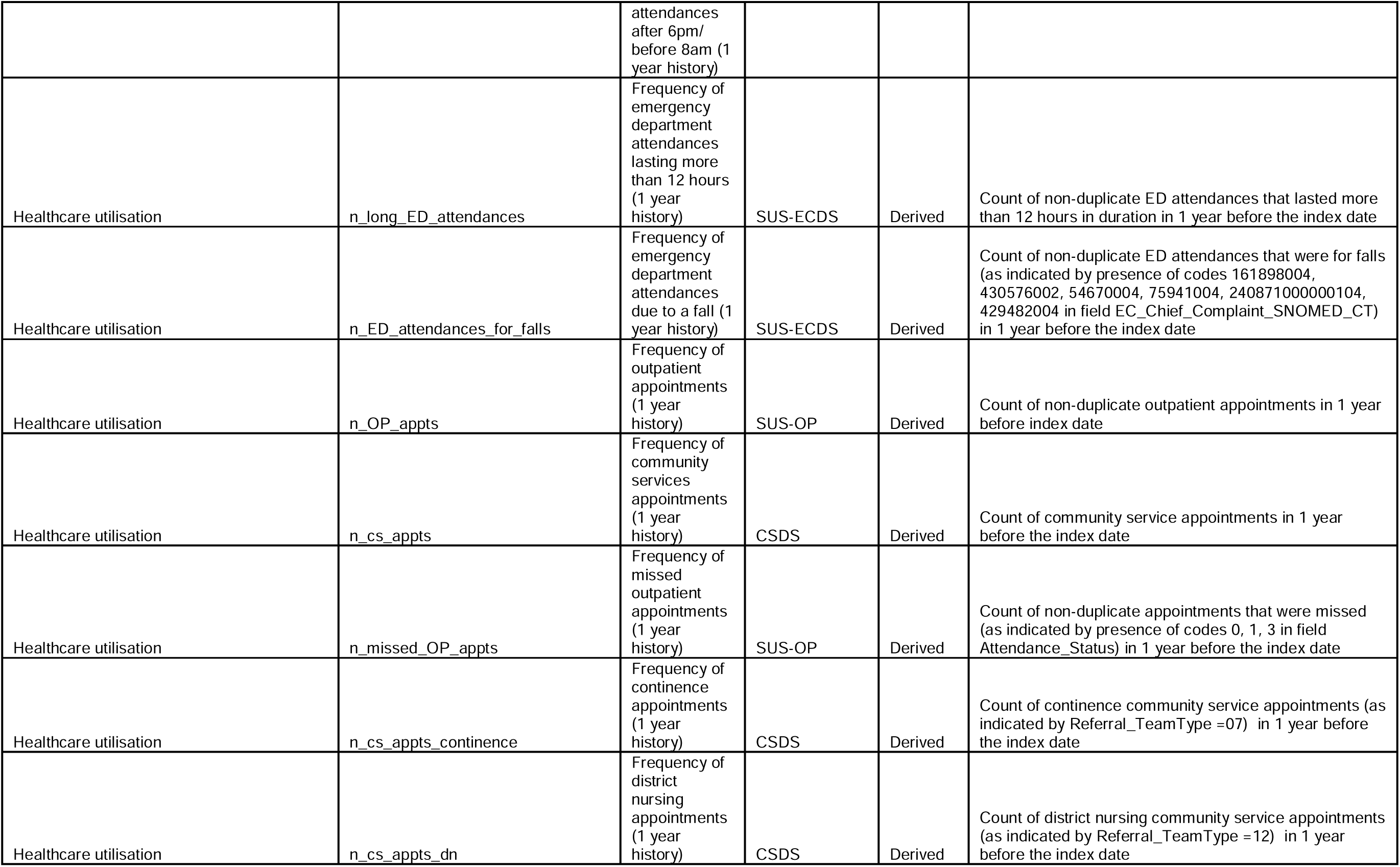
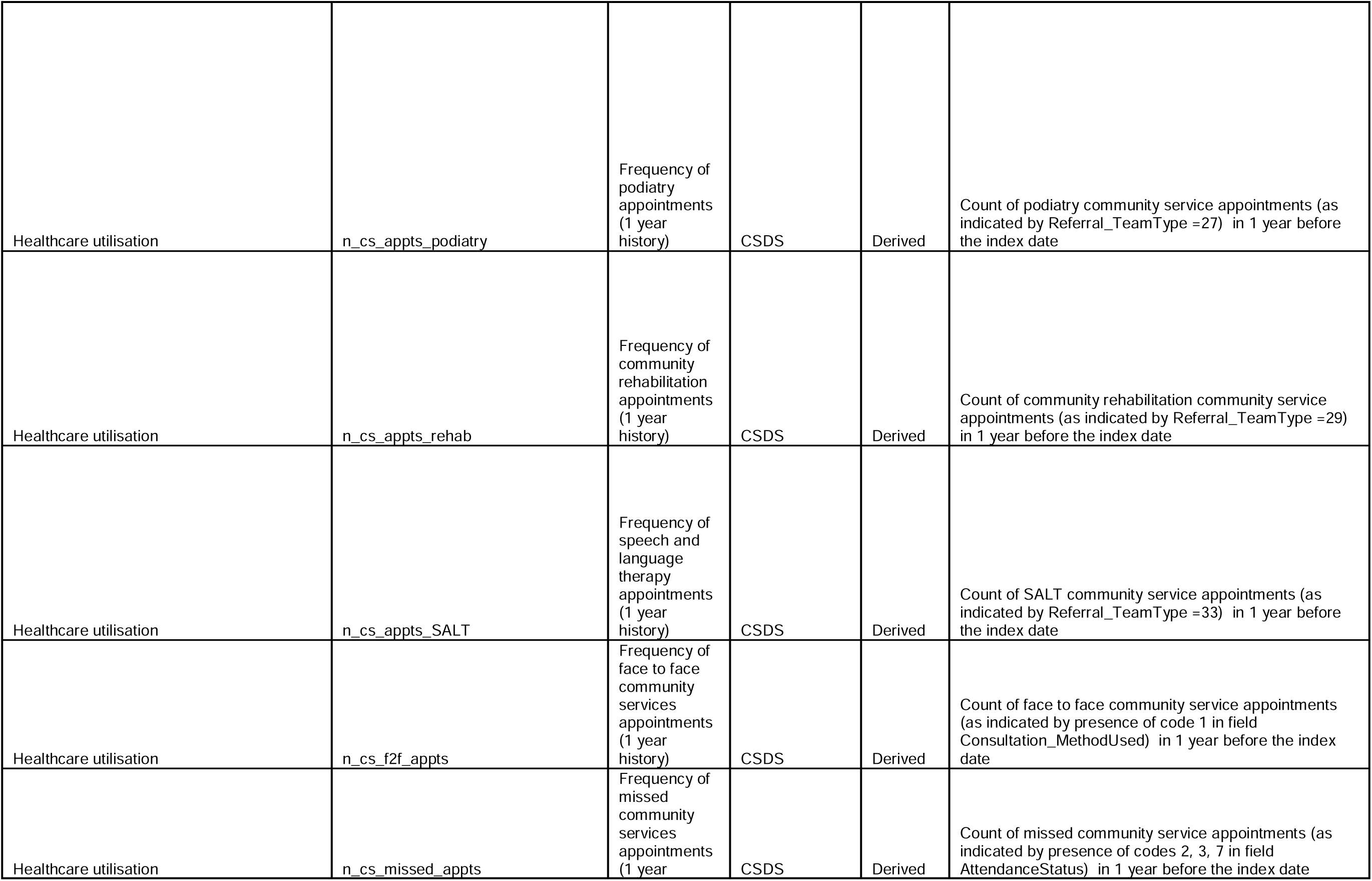
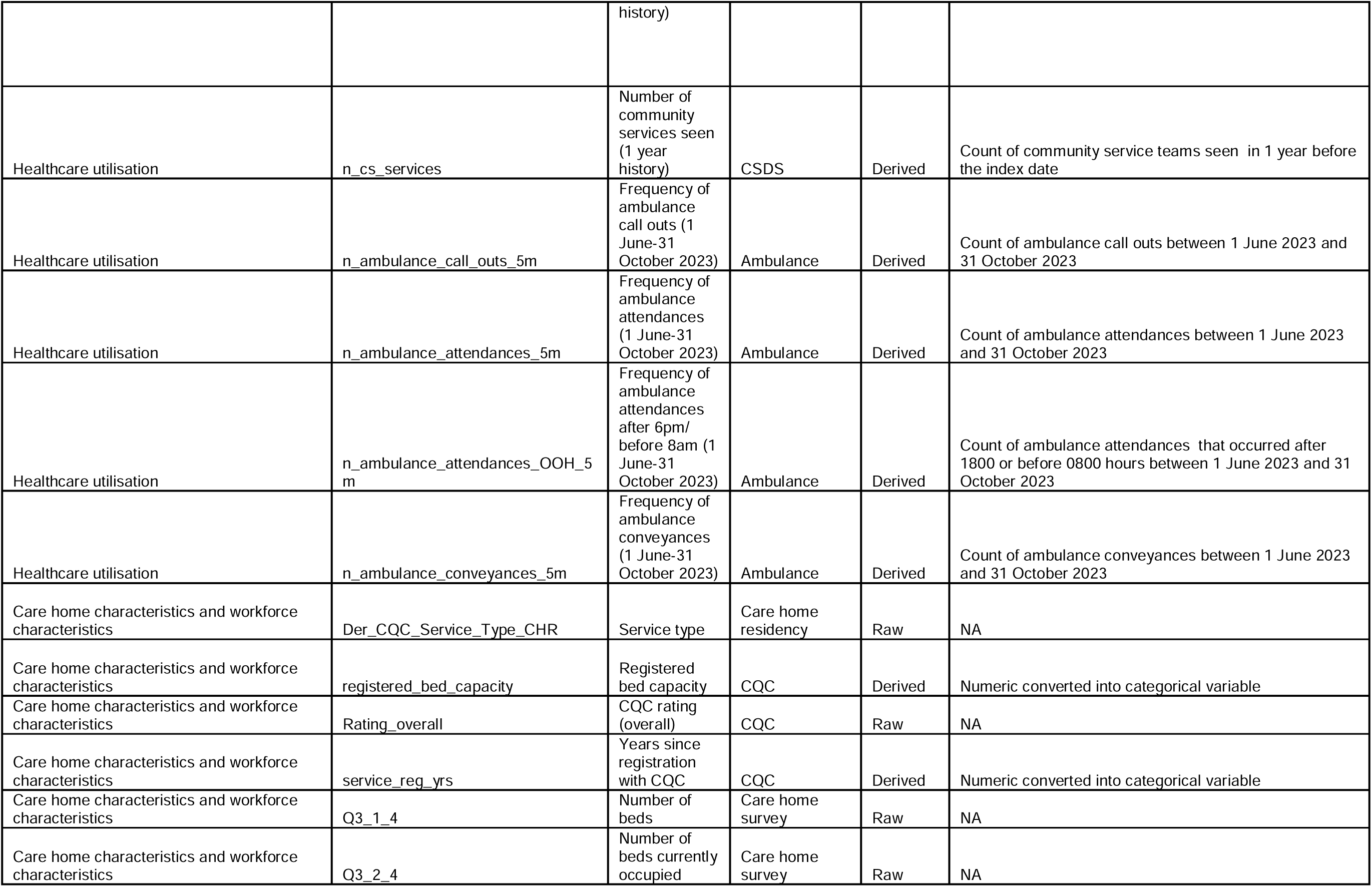
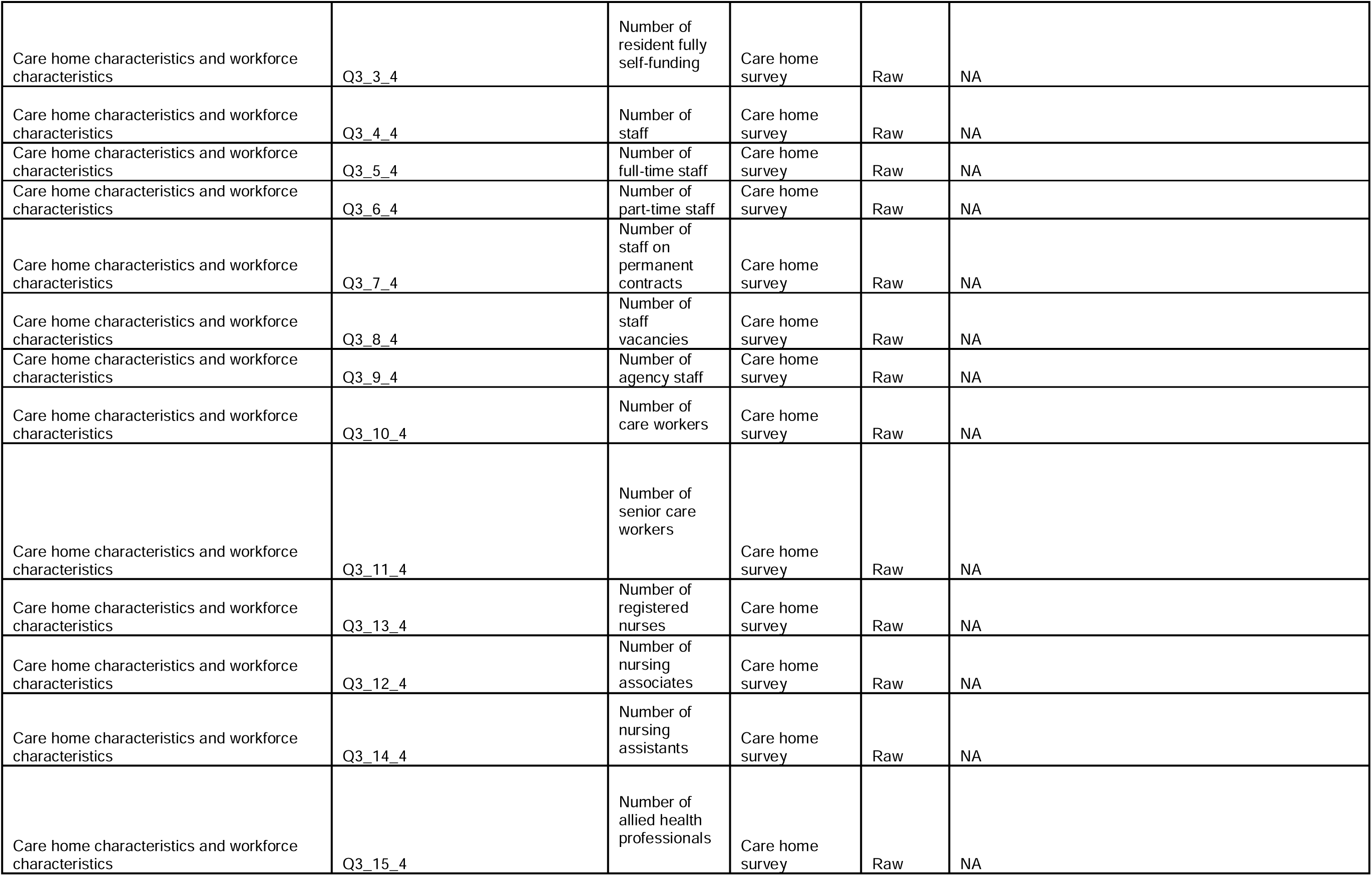
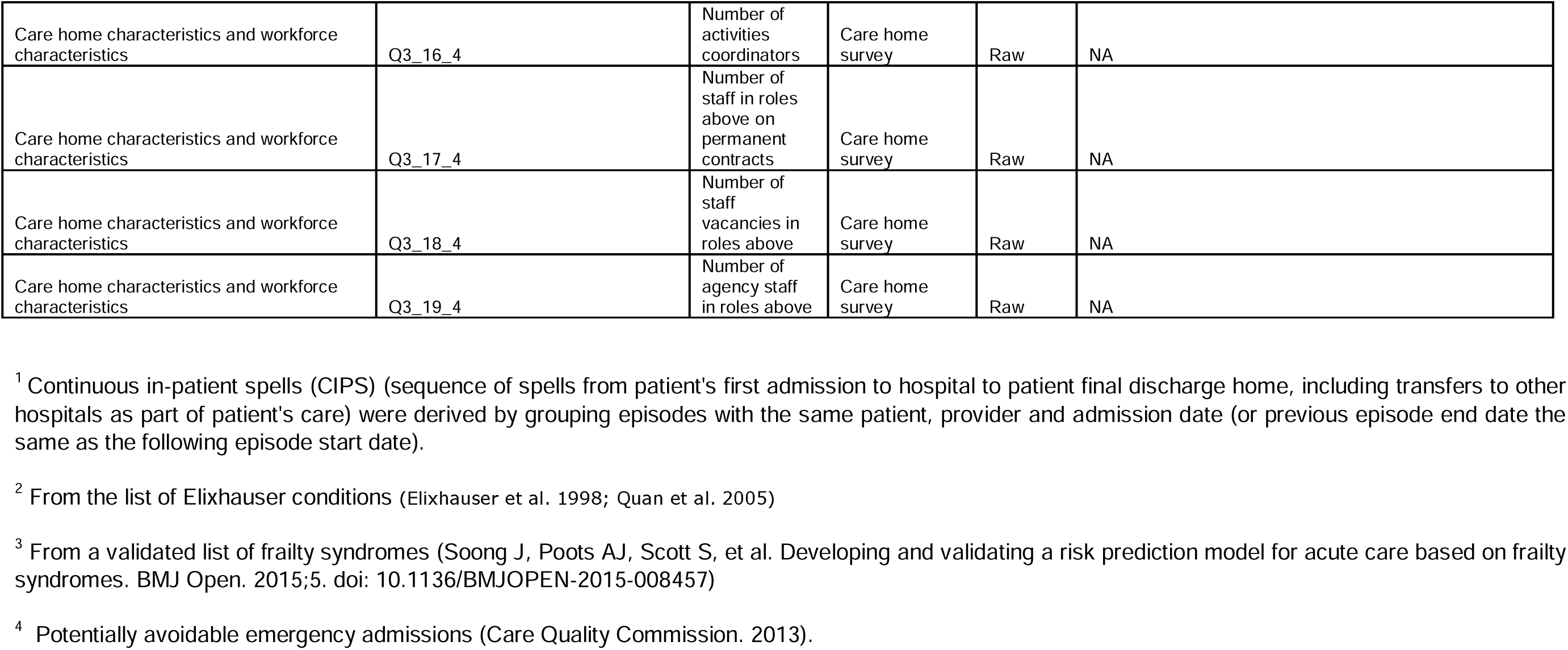
Table a: Data specification for final prototype MDS

Table b: data specification for cleaned datasets

**Table b: data specification for cleaned datasets**
| Dataset | Domain | Variable name | Description |
| --- | --- | --- | --- |
| ALL | Linkage | pseudonhsno | Pseudonymised NHS number |
| SUS-APCE | Healthcare utilization | Generated_Record_ID | Generated Record ID |
| SUS-APCE | Healthcare utilization | Der_Postcode_LSOA_Code | LSOA code |
| SUS-APCE | Healthcare utilization | GP_Practice_Code | GP practice code |
| SUS-APCE | Healthcare utilization | Der_Postcode_CCG_Code | CCG code |
| SUS-APCE | Healthcare utilization | Der_Financial_Year | Financial year |
| SUS-APCE | Healthcare utilization | Der_Activity_Month | Activity month |
| SUS-APCE | Healthcare utilization | Admission_Date | Admission date |
| SUS-APCE | Healthcare utilization | Admission_Time | Admission time |
| SUS-APCE | Healthcare utilization | Admission_Method | Admission method |
| SUS-APCE | Healthcare utilization | Source_of_Admission | Source of admission |
| SUS-APCE | Healthcare utilization | Administrative_Category | Administrative category |
| SUS-APCE | Healthcare utilization | Discharge_Date | Discharge date |
| SUS-APCE | Healthcare utilization | Discharge_Time | Discharge time |
| SUS-APCE | Healthcare utilization | Discharge_Method | Discharge method |
| SUS-APCE | Healthcare utilization | Discharge_Destination | Discharge destination |
| SUS-APCE | Healthcare utilization | Der_Provider_Code | Provider code |
| SUS-APCE | Diagnoses | Der_Diagnosis_Count | Count of diagnosis codes |
| SUS-APCE | Diagnoses | Der_Primary_Diagnosis_Code | Primary diagnosis code |
| SUS-APCE | Diagnoses | Der_Secondary_Diagnosis_Code_1 | Secondary diagnosis code 1 |
| SUS-APCE | Diagnoses | Der_Secondary_Diagnosis_Code_2 | Secondary Diagnosis Code 2 |
| SUS-APCE | Diagnoses | Der_Secondary_Diagnosis_Code_3 | Secondary Diagnosis Code 3 |
| SUS-APCE | Diagnoses | Der_Secondary_Diagnosis_Code_4 | Secondary Diagnosis Code 4 |
| SUS-APCE | Diagnoses | Der_Secondary_Diagnosis_Code_5 | Secondary Diagnosis Code 5 |
| SUS-APCE | Diagnoses | Der_Secondary_Diagnosis_Code_6 | Secondary Diagnosis Code 6 |
| SUS-APCE | Diagnoses | Der_Secondary_Diagnosis_Code_7 | Secondary Diagnosis Code 7 |
| SUS-APCE | Diagnoses | Der_Secondary_Diagnosis_Code_8 | Secondary Diagnosis Code 8 |
| SUS-APCE | Diagnoses | Der_Secondary_Diagnosis_Code_9 | Secondary Diagnosis Code 9 |
| SUS-APCE | Diagnoses | Der_Secondary_Diagnosis_Code_10 | Secondary Diagnosis Code 10 |
| SUS-APCE | Diagnoses | Der_Secondary_Diagnosis_Code_11 | Secondary Diagnosis Code 11 |
| SUS-APCE | Diagnoses | Der_Secondary_Diagnosis_Code_12 | Secondary Diagnosis Code 12 |
| SUS-APCE | Diagnoses | Der_Secondary_Diagnosis_Code_13 | Secondary Diagnosis Code 13 |
| SUS-APCE | Diagnoses | Der_Secondary_Diagnosis_Code_14 | Secondary Diagnosis Code 14 |
| SUS-APCE | Diagnoses | Der_Secondary_Diagnosis_Code_15 | Secondary Diagnosis Code 15 |
| SUS-APCE | Diagnoses | Der_Secondary_Diagnosis_Code_16 | Secondary Diagnosis Code 16 |
| SUS-APCE | Diagnoses | Der_Secondary_Diagnosis_Code_17 | Secondary Diagnosis Code 17 |
| SUS-APCE | Diagnoses | Der_Secondary_Diagnosis_Code_18 | Secondary Diagnosis Code 18 |
| SUS-APCE | Diagnoses | Der_Secondary_Diagnosis_Code_19 | Secondary Diagnosis Code 19 |
| SUS-APCE | Diagnoses | Der_Secondary_Diagnosis_Code_20 | Secondary Diagnosis Code 20 |
| SUS-APCE | Diagnoses | Der_Secondary_Diagnosis_Code_21 | Secondary Diagnosis Code 21 |
| SUS-APCE | Diagnoses | Der_Secondary_Diagnosis_Code_22 | Secondary Diagnosis Code 22 |
| SUS-APCE | Diagnoses | Der_Secondary_Diagnosis_Code_23 | Secondary Diagnosis Code 23 |
| SUS-APCE | Diagnoses | Der_Diagnosis_All | All diagnosis codes |
| SUS-APCE | Healthcare utilization | Treatment_Function_Code | Treatment function code |
| SUS-APCE | Healthcare utilization | Main_Speciality_Code | Main speciality code |
| SUS-APCE | Healthcare utilization | Patient_Classification | Patient classification |
| SUS-APCE | Healthcare utilization | Episode_Start_Date | Episode Start Date |
| SUS-APCE | Healthcare utilization | Episode_End_Date | Episode End Date |
| SUS-APCE | Healthcare utilization | Der_Episode_Number | Episode Number |
| SUS-APCE | Healthcare utilization | Der_Spell_ID | Spell ID |
| SUS-APCE | Demographics/characteristics | Month_of_Birth_SUS | Month of birth |
| SUS-APCE | Demographics/characteristics | Year_of_Birth_SUS | Year of birth |
| SUS-APCE | Demographics/characteristics | Sex | Sex |
| SUS-APCE | Demographics/characteristics | Ethnic_Group | Ethnicity |
| SUS-ECDS | Healthcare utilization | Arrival_Date | Date of arrival |
| SUS-ECDS | Healthcare utilization | Arrival_Time | Time of arrival |
| SUS-ECDS | Healthcare utilization | EC_Treatment_Date_01 | Date of treatment |
| SUS-ECDS | Healthcare utilization | EC_Seen_For_Treatment_Time_Since_Arrival | Waiting time from arrival to being seen for treatment |
| SUS-ECDS | Healthcare utilization | EC_Departure_Date | Date of departure |
| SUS-ECDS | Healthcare utilization | EC_Departure_Time | Time of departure |
| SUS-ECDS | Healthcare utilization | Der_EC_Duration | Duration of the A&E attendance |
| SUS-ECDS | Healthcare utilization | Der_Activity_Month | Activity month |
| SUS-ECDS | Healthcare utilization | Generated_Record_ID | Generated Record ID |
| SUS-ECDS | Healthcare utilization | Provider_Code | Provider code |
| SUS-ECDS | Healthcare utilization | EC_Department_Type | Department type |
| SUS-ECDS | Healthcare utilization | Der_Postcode_LSOA_2011_Code | LSOA code |
| SUS-ECDS | Healthcare utilization | Der_Postcode_CCG_Code | CCG code |
| SUS-ECDS | Healthcare utilization | GP_Practice_Code | GP practice code |
| SUS-ECDS | Healthcare utilization | Accommodation_Status_SNOMED_CT | Accommodation status (SNOMED CT) |
| SUS-ECDS | Healthcare utilization | EC_AttendanceCategory | Attendance category (SNOMED CT) |
| SUS-ECDS | Healthcare utilization | EC_Arrival_Mode_SNOMED_CT | Arrival mode (SNOMED CT) |
| SUS-ECDS | Healthcare utilization | EC_Attendance_Number | Attendance number (SNOMED CT) |
| SUS-ECDS | Healthcare utilization | Discharge_Destination_SNOMED_CT | Discharge destination (SNOMED CT) |
| SUS-ECDS | Healthcare utilization | Discharge_Follow_Up_SNOMED_CT | Discharge follow up (SNOMED CT) |
| SUS-ECDS | Healthcare utilization | EC_Acuity_SNOMED_CT | Acuity (SNOMED CT) |
| SUS-ECDS | Healthcare utilization | EC_Attendance_Source_SNOMED_CT | Attendance source (SNOMED CT) |
| SUS-ECDS | Healthcare utilization | EC_Discharge_Status_SNOMED_CT | Discharge status (SNOMED CT) |
| SUS-ECDS | Complications/adverse events | EC_Chief_Complaint_SNOMED_CT | Chief Complaint (SNOMED CT) |
| SUS-ECDS | Complications/adverse events | EC_Diagnosis_01 | Primary diagnosis (SNOMED CT) |
| SUS-ECDS | Healthcare utilization | EC_Treatment_01 | Primary treatment code (SNOMED CT) |
| SUS-ECDS | Complications/adverse events | Der_EC_Diagnosis_All | All diagnosis codes (SNOMED CT) |
| SUS-ECDS | Resident needs | Der_EC_Treatment_All | All treatment codes (SNOMED CT) |
| SUS-ECDS | Complications/adverse events | EC_Injury_Date | Injury date |
| SUS-ECDS | Complications/adverse events | EC_Injury_Time | Injury time |
| SUS-ECDS | Complications/adverse events | EC_Injury_Activity_Status_SNOMED | Injury activity status (SNOMED CT) |
| SUS-ECDS | Complications/adverse events | EC_Injury_Activity_Type_SNOMED_C | Injury activity type (SNOMED CT) |
| SUS-ECDS | Complications/adverse events | EC_Injury_Intent_SNOMED_CT | Injury intent (SNOMED CT) |
| SUS-ECDS | Complications/adverse events | EC_Injury_Mechanism_SNOMED_CT | Injury mechanism (SNOMED CT) |
| SUS-ECDS | Complications/adverse events | EC_Place_Of_Injury_SNOMED_CT | Place of injury (SNOMED CT) |
| SUS-ECDS | Complications/adverse events | AEA_Diagnosis_01 | Primary diagnosis code (AEA code) |
| SUS-ECDS | Healthcare utilization | AEA_Treatment_01 | Primary treatment code (AEA code) |
| SUS-ECDS | Complications/adverse events | Der_AEA_Diagnosis_All | All diagnosis codes (AEA codes) |
| SUS-ECDS | Resident needs | Der_AEA_Treatment_All | All treatment codes (AEA codes) |
| SUS-ECDS | Healthcare utilization | Org_Code_Patient_Pathway_ID_Issuer | Organisation code of the organisation that assigned the patient pathway identifier |
| SUS-ECDS | Demographics/characteristics | Month_of_Birth | Month of birth |
| SUS-ECDS | Demographics/characteristics | Year_of_Birth | Year of birth |
| SUS-ECDS | Demographics/characteristics | Sex | Sex |
| SUS-ECDS | Demographics/characteristics | Ethnic_Category | Ethnicity |
| SUS-OP | Healthcare utilization | Appointment_Date | Appointment date |
| SUS-OP | Healthcare utilization | Appointment_Time | Appointment time |
| SUS-OP | Healthcare utilization | Der_Activity_Month | Activity month |
| SUS-OP | Healthcare utilization | Der_Financial_Year | Financial year |
| SUS-OP | Healthcare utilization | Der_Provider_Code | Provider code |
| SUS-OP | Healthcare utilization | Der_Postcode_LSOA_Code | LSOA code |
| SUS-OP | Healthcare utilization | Der_Postcode_CCG_Code | CCG code |
| SUS-OP | Healthcare utilization | GP_Practice_Code | GP practice code |
| SUS-OP | Healthcare utilization | Der_Appointment_Type | Appointment type |
| SUS-OP | Healthcare utilization | Der_Attendance_Type | Attendance type |
| SUS-OP | Healthcare utilization | Attendance_Status | Attendance status |
| SUS-OP | Healthcare utilization | OPA_Referral_Source | Source of referral for the outpatient episode |
| SUS-OP | Healthcare utilization | Treatment_Function_Code | Treatment function code |
| SUS-OP | Healthcare utilization | Main_Speciality_Code | Main speciality code |
| SUS-OP | Resident needs | Der_Procedure_All | All procedure codes |
| SUS-OP | Demographics/characteristics | Month_of_Birth_SUS | Month of birth |
| SUS-OP | Demographics/characteristics | Year_of_Birth_SUS | Year of birth |
| SUS-OP | Demographics/characteristics | Sex | Sex |
| SUS-OP | Demographics/characteristics | Ethnic_Category | Ethnicity |
| PDS | Demographics/characteristics | Der_Practice_Code | GP practice code |
| PDS | Demographics/characteristics | Der_CCGofResidence | CCG code for residence location |
| PDS | Demographics/characteristics | Gender | Sex |
| PDS | Demographics/characteristics | DateofDeath | Date of death |
| PDS | Demographics/characteristics | Der_Postcode_LSOA_code | LSOA of residence |
| PDS | Demographics/characteristics | Der_DOBYearMnth | Year and month of birth |
| PDS | Demographics/characteristics | Change_Time_Stamp | Date and time to indicate when record was updated (for any column) |
| Care home residency<sup>1</sup> and CQC dataset | Linkage | Der_CQC_Location | Pseudonymised CQC location id |
| Care home residency | Care home stay | Der_Start_Date | Start of care home stay |
| Care home residency | Care home stay | Der_End_Date | End of care home stay |
| Care home residency | Care home characteristics and workforce characteristics | Der_CQC_Service_Type | Type of care home |
| Care home residency | Demographics/characteristics | CCG_Of_Residence | CCG of care home |
| Care home residency | Care home characteristics and workforce characteristics | CareHomeIndicator | ? Unclear |
| Care home residency | Care home stay | DeathFlag | Flag to indicate resident died |
| Care home residency | Care home stay | YearMonthDeath_YYYYMM | Year and month of death |
| Care home residency | Care home stay | DeathInHospFlag | Flag to indicate death in hospital |
| Care home residency | Care home stay | LastResidencyFlag | Flag to indicate last residence of the person |
| Ambulance | Healthcare utilization | call_date | Date of call |
| Ambulance | Healthcare utilization | call_origin | Codes to identify the origin of the call into the ambulance control room, whether 999 was directly dialled or it was transferred from another agency. |
| Ambulance | Healthcare utilization | stop_codes | Codes to identify the reason the call was closed if no face to face response was received from the ambulance service. |
| Ambulance | Healthcare utilization | time_call_connected | Time call connected |
| Ambulance | Healthcare utilization | time_call_answered | Time call answered |
| Ambulance | Healthcare utilization | treatment_type | Codes to identify the overall outcome of the call. |
| Ambulance | Healthcare utilization | chief_complaint_call_triage_code | Codes to identify the initial chief complaint of the patient based on the information provided during telephone call to the ambulance control room. |
| Ambulance | Healthcare utilization | orgid_prov3 | Provider code 3 characters |
| Ambulance | Healthcare utilization | time_resource_arrived_on_scene | Time resource arrived on scene |
| Ambulance | Healthcare utilization | receiving_location_type_cad | ? Unclear |
| ASC CLD | Care home stay | Accommodation_Status_ASC | Accommodation status |
| ASC CLD | Care home stay | Client_Funding_Status_ASC | Client funding status |
| ASC CLD | Care home stay | Service_Component_ASC | Service component |
| ASC CLD | Care home stay | Service_Type_ASC | Service type |
| CSDS (CYP001MPI) | Demographics/characteristics | Gender | Person stated gender code or Sex |
| CSDS (CYP001MPI) | Demographics/characteristics | EthnicCategory | Ethnic category |
| CSDS (all files) | Healthcare utilization | EFFECTIVE_FROM | Date and time to indicate when record was updated (for any column) |
| CSDS (all files) | Healthcare utilization | RecordNumber | Record number |
| CSDS (CYP001MPI) | Healthcare utilization | LSOA | LSOA code |
| CSDS (CYP001MPI) | Healthcare utilization | Der_Postcode_yr2011_LSOA | LSOA code |
| CSDS (CYP001MPI) | Demographics/characteristics | AgeYr_RP_StartDate | Age of patient at reporting period start (days) |
| CSDS (CYP001MPI) | Care home stay | AgeYr_Death | Age at death (years) |
| CSDS (CYP001MPI) | Care home stay | Age_Death | Age at death (days) |
| CSDS (CYP001MPI) | Healthcare utilization | OrgIDICBRes | Organisation identifier (ICB of residence) |
| CSDS (CYP001MPI) | Palliative care needs | DiscussPreferredDeathLocation_Indicator | Preferred death location discussed indicator |
| CSDS (CYP001MPI) | Palliative care needs | DeathLocationPreferred_Type | Death location type code (preferred) |
| CSDS (CYP001MPI) | Palliative care needs | DeathLocationActual_Type | Death location type code (actual) |
| CSDS (CYP001MPI) | Palliative care needs | NotAtPreferredLocation_Reason | Death not at preferred location reason |
| CSDS (CYP201CareContact) | Healthcare utilization | CareContactID | Care contact identifier |
| CSDS (CYP201CareContact) | Healthcare utilization | ServiceRequestID | Service request identifier |
| CSDS (CYP201CareContact) | Healthcare utilization | Contact_Date | Care contact date |
| CSDS (CYP201CareContact) | Healthcare utilization | Consultation_Type | Consultation type |
| CSDS (CYP201CareContact) | Healthcare utilization | Consultation_MediumUsed | Consultation mechanism e.g. face to face |
| CSDS (CYP201CareContact) | Healthcare utilization | Activity_LocationType | Activity location type code |
| CSDS (CYP201CareContact) | Healthcare utilization | OrgID_Provider | Organisation identifier (code of provider) |
| CSDS (CYP201CareContact) | Healthcare utilization | AgeYr_Contact_Date | Age at care contact date |
| CSDS (CYP201CareContact) | Healthcare utilization | AttendanceStatus | Attendance status |
| CSDS (CYP101Referral) | Healthcare utilization | SourceOfReferral | Source of referral |
| CSDS (CYP101Referral) | Healthcare utilization | Referring_StaffGroup | Referring care professional staff group (community care) |
| CSDS (CYP101Referral) | Healthcare utilization | PrimaryReferralReason | Primary reason for referral (community care) |
| CSDS (CYP102ServiceTypeReferredTo) | Healthcare utilization | ServiceRequestID | Service request identifier |
| CSDS (CYP102ServiceTypeReferredTo) | Healthcare utilization | TeamID_Local | Care professional team local identifier |
| CSDS | Healthcare utilization | TeamType | Service or team type referred to (community care) |
| (CYP102ServiceTypeReferredTo) |  |  |  |
| CQC | Care home characteristics and workforce characteristics | Location_HSCA_start_date | Start of registration at location |
| CQC | Care home characteristics and workforce characteristics | Care_home? | Care home or not |
| CQC | Care home characteristics and workforce characteristics | Care_homes_beds | Number of beds |
| CQC | Care home characteristics and workforce characteristics | Location_Inspection_Directorate | Inspection Directorate |
| CQC | Care home characteristics and workforce characteristics | Location_Primary_Inspection_Cat e | Primary inspection category |
| CQC | Care home characteristics and workforce characteristics | Publication_Date | Publication date |
| CQC | Care home characteristics and workforce characteristics | Location_Region | Region |
| CQC | Care home characteristics and workforce characteristics | Location_NHS_Region | NHS Region |
| CQC | Care home characteristics and workforce characteristics | Location_Local_Authority | Location Authority |
| CQC | Care home characteristics and workforce characteristics | Location_ONSPD_CCG | CCG |
| CQC | Care home characteristics and workforce characteristics | service_type | Service type e.g. nursing home |
| CQC | Care home characteristics and workforce characteristics | Location_Type | Location type |
| CQC | Care home characteristics and workforce characteristics | Rating_overall | Rating: overall |
| CQC | Care home characteristics and workforce characteristics | Rating_caring | Rating: caring |
| CQC | Care home characteristics and workforce characteristics | Rating_well_led | Rating: well led |
| CQC | Care home characteristics and workforce characteristics | Rating_effective | Rating: effective |
| CQC | Care home characteristics and workforce characteristics | Rating_reponsive | Rating: responsive |
| CQC | Care home characteristics and workforce characteristics | Rating_safe | Rating: safe |
| CQC | Care home characteristics and workforce characteristics | serv_user_dementia | Service users with dementia |
| CQC | Care home characteristics and workforce characteristics | years_since_registration | Years since registration |

## APPENDIX 6 Completion of measures added into care home digital care record software, by wave, for linked DCR data*

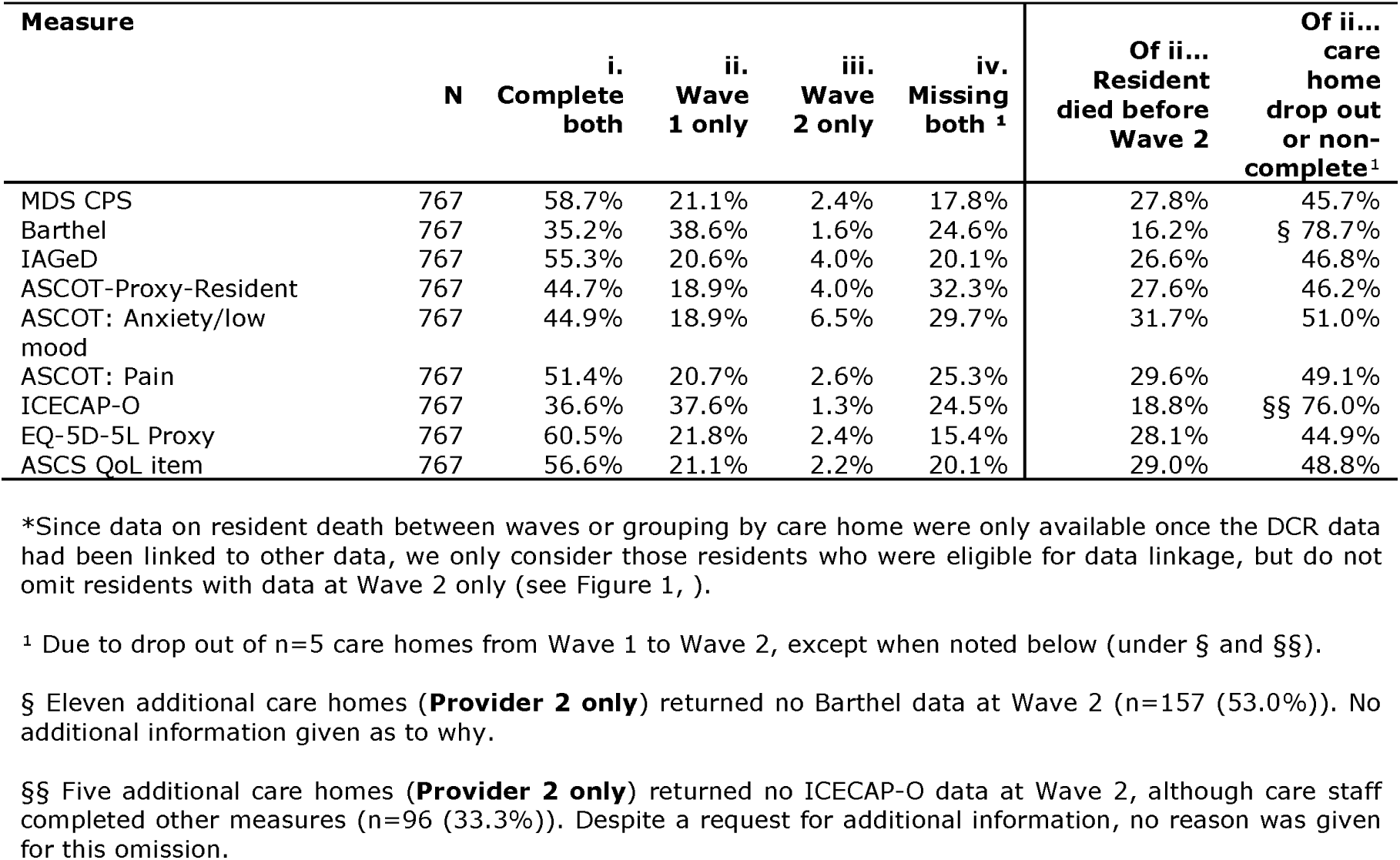

| Measure | N | i.<br>Complete<br>both | ii.<br>Wave<br>1 only | iii.<br>Wave<br>2 only | iv.<br>Missing<br>both <sup>1</sup> | Of ii...<br>Resident<br>died before<br>Wave 2 | Of ii...<br>care<br>home<br>drop out<br>or non-<br>complete<sup>1</sup> |
| --- | --- | --- | --- | --- | --- | --- | --- |
| MDS CPS | 767 | 58.7% | 21.1% | 2.4% | 17.8% | 27.8% | 45.7% |
| Barthel | 767 | 35.2% | 38.6% | 1.6% | 24.6% | 16.2% | § 78.7% |
| IAGeD | 767 | 55.3% | 20.6% | 4.0% | 20.1% | 26.6% | 46.8% |
| ASCOT-Proxy-Resident | 767 | 44.7% | 18.9% | 4.0% | 32.3% | 27.6% | 46.2% |
| ASCOT: Anxiety/low<br>mood | 767 | 44.9% | 18.9% | 6.5% | 29.7% | 31.7% | 51.0% |
| ASCOT: Pain | 767 | 51.4% | 20.7% | 2.6% | 25.3% | 29.6% | 49.1% |
| ICECAP-O | 767 | 36.6% | 37.6% | 1.3% | 24.5% | 18.8% | §§ 76.0% |
| EQ-5D-5L Proxy | 767 | 60.5% | 21.8% | 2.4% | 15.4% | 28.1% | 44.9% |
| ASCS QoL item | 767 | 56.6% | 21.1% | 2.2% | 20.1% | 29.0% | 48.8% |

\*Since data on resident death between waves or grouping by care home were only available once the DCR data had been linked to other data, we only consider those residents who were eligible for data linkage, but do not omit residents with data at Wave 2 only (see Figure 1, ).
<sup>1</sup> Due to drop out of n=5 care homes from Wave 1 to Wave 2, except when noted below (under § and §§).
§ Eleven additional care homes (**Provider 2 only**) returned no Barthel data at Wave 2 (n=157 (53.0%)). No additional information given as to why.
§§ Five additional care homes (**Provider 2 only**) returned no ICECAP-O data at Wave 2, although care staff completed other measures (n=96 (33.3%)). Despite a request for additional information, no reason was given for this omission.

## APPENDIX 7: GP data items possible to access from one ICS

| Type | Variable |
| --- | --- |
| Demographics | Ethnicity |
| Demographics | BMI |
| Demographics | Age in Years |
| Demographics | Has died Y/N /month and year of death |
| Demographics | Care home flag in GP record and/ or ICS system |
| Demographics | Registration with GP practice aligned to care home declined |
| Risk stratification | End-of-life pathway register |
| Risk stratification | Place of death discussed |
| Risk stratification | Preferred place of death |
| Risk stratification | Infection - urinary tract |
| Risk stratification | Infection - chest - lower respiratory tract infection |
| Risk stratification | Infection - skin |
| Risk stratification | Injury resulting from fall |
| Risk stratification | Haematological malignancies |
| Risk stratification | Depression |
| Risk stratification | Dementia |
| Risk stratification | Incontinence - fecal |
| Risk stratification | Delirium |
| Risk stratification | Difficulty swallowing |
| Risk stratification | Frailty index (eFi) |
| Risk stratification | eFI: activity limitation |
| Risk stratification | eFI: anaemia and haematinic deficiency |
| Risk stratification | eFI: arthritis |
| Risk stratification | eFI: atrial fibrillation |
| Risk stratification | eFI: cerebrovascular disease |
| Risk stratification | eFI: chronic kidney disease (CKD) |
| Risk stratification | eFI: diabetes |
| Risk stratification | eFI: dizziness |
| Risk stratification | eFI: dyspnoea |
| Risk stratification | eFI: falls |
| Risk stratification | eFI: foot problems |
| Risk stratification | eFI: fragility fracture |
| Risk stratification | eFI: hearing impairment |
| Risk stratification | eFI: heart failure |
| Risk stratification | eFI: heart valve disease |
| Risk stratification | eFI: housebound |
| Risk stratification | eFI: hypertension |
| Risk stratification | eFI: hypotension/syncope |
| Risk stratification | eFI Memory & cognitive problems |
| Risk stratification | eFI Mobility and transfer problems |
| Risk stratification | eFI: osteoporosis |
| Risk stratification | eFI: Parkinsonism and tremor |
| Risk stratification | eFI: peptic ulcer |
| Risk stratification | eFI: Peripheral vascular disease |
| Risk stratification | eFI: polypharmacy |
| Risk stratification | eFI: requirement for care |
| Risk stratification | eFI: respiratory disease |
| Risk stratification | eFI: skin ulcer |
| Risk stratification | eFI: sleep disturbance |
| Risk stratification | eFI: social vulnerability |
| Risk stratification | eFI: thyroid disease |
| Risk stratification | eFI: urinary incontinence |
| Risk stratification | eFI: urinary system disease |
| Risk stratification | eFI: visual impairment |
| Risk stratification | eFI: weight loss and anorexia |
| Risk stratification | eFI: ischaemic heart disease |
| Medications | Numbers of current prescriptions |
| Medications | Most recent medication review |
| Medications | Penicillins (BNF 5.1.1) |
| Medications | Cephalosporins, carbapenems & other beta-lactams (BNF 5.1.2) |
| Medications | Tetracyclines (BNF 5.1.3) |
| Medications | Aminoglycosides (BNF 5.1.4) |
| Medications | Macrolides (BNF 5.1.5) |
| Medications | Sulfonamides and trimethoprim (BNF 5.1.8) |
| Medications | Quinolones (BNF 5.1.12) |
| Medications | Non-opioid analgesics (BNF 4.7.1) |
| Medications | Compound analgesic preparations (BNF 4.7.1) |
| Medications | Opioid analgesics (BNF 4.7.2) |
| Medications | Non-steroidal anti-inflammatory drugs (BNF 10.1.1) |
| Medications | Other drugs used as analgesics |
| Vaccinations | Flu |
| Vaccinations | Covid 19 |
| Vaccinations | Pneumonia |
| Appointments/utilisation | Number of appointments by staff type |
| Appointments/utilisation | EHCH MDT referral |
| Appointments/utilisation | Hospital referral recorded? |
| Appointments/utilisation | A&E referral recorded? |
| Appointments/utilisation | Continence service referral recorded? |
| Appointments/utilisation | Community services referral recorded? |
| Appointments/utilisation | Community nurse referral recorded |
| Appointments/utilisation | Referrals to falls service |
| Appointments/utilisation | Face to Face |
| Appointments/utilisation | Telephone appointments |

## APPENDIX 8 Reason for non-inclusion of planned variables in final MDS

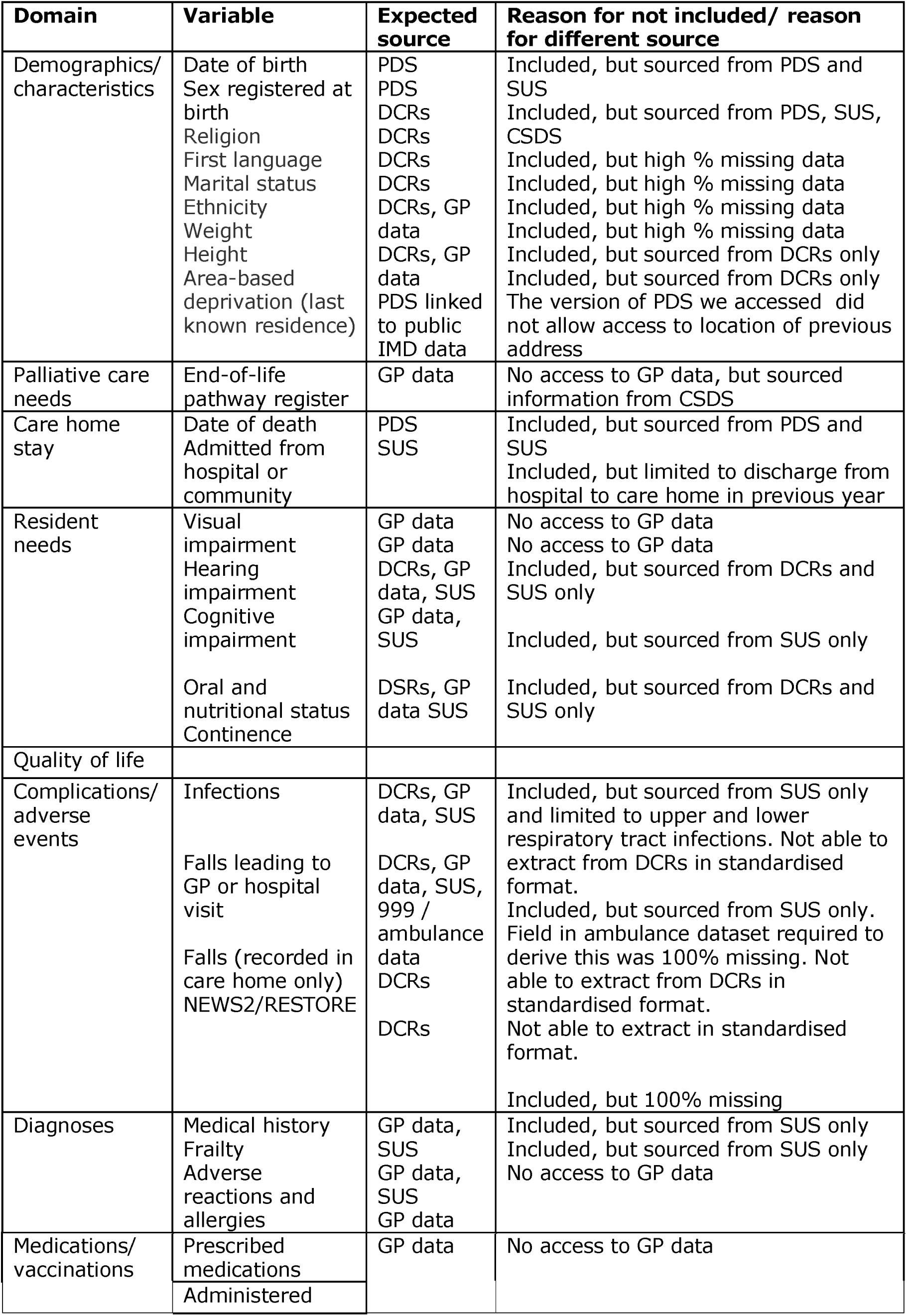
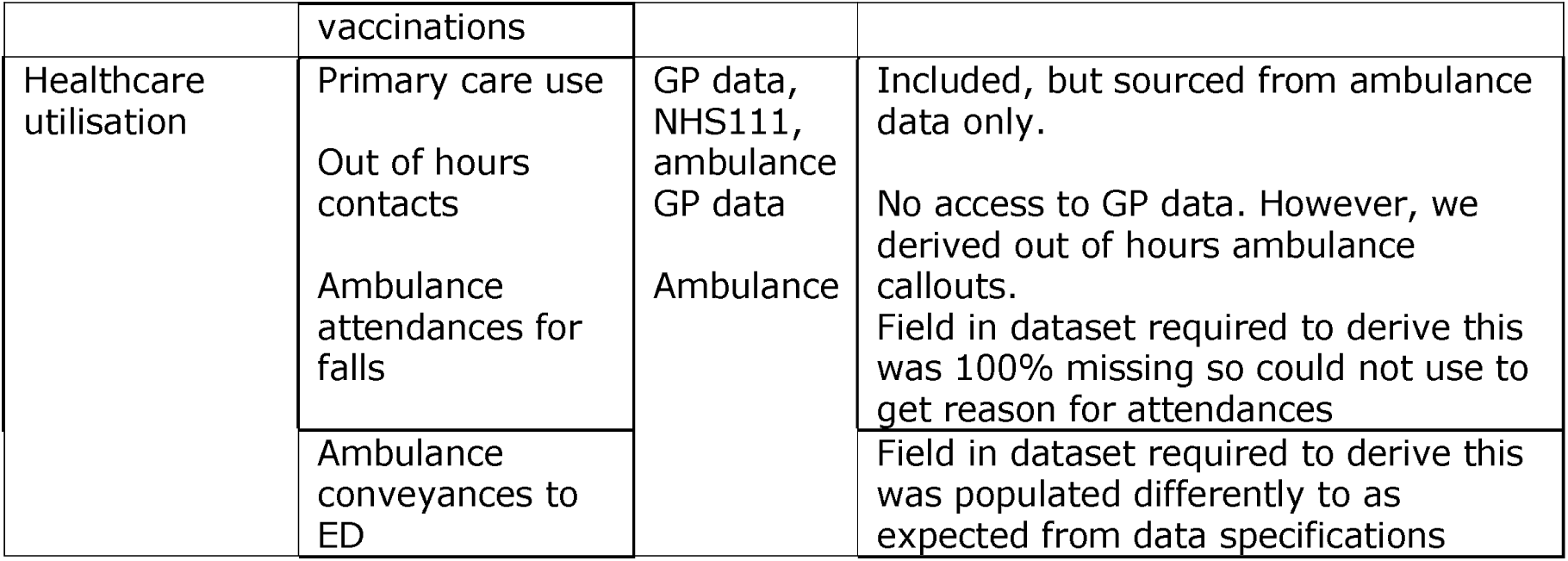

## APPENDIX 9 Comparison of variables with inconsistent definitions across data sources

A small number of comorbidities were recorded in both hospital and care home records. However, these variables were not defined in a consistent way. Even within a data source, there can be several established definitions and code lists, which will give slightly different results. Table a shows that 11% of residents in our sample are identified as having dementia from SUS data according to one definition but not another (53 + 13/583).

The comorbidities were also collected differently: variables in the care home data were recorded at one particular point in time - therefore reflecting the resident’s health status at that moment. For the SUS data, we collected information relating to these conditions over a three-year look-back period. For acute conditions, there were high levels of discrepancy between SUS and care home DCRs. For example, delirium was recorded in hospital records for 145 patients in the previous 3 years, but only 20 (14%) of these were recorded as delirious by care home staff at the time of recording. However, there were also 30 (48%, 30/62) residents recorded with delirium in the care home DCRs that were not reflected in hospital records prior to that date. For cognitive impairment, which tends to not improve over time, there was more consistency, although still substantial disagreement. Agreement between care home DCR and SUS record of dementia is recorded in Table 3 in main results.

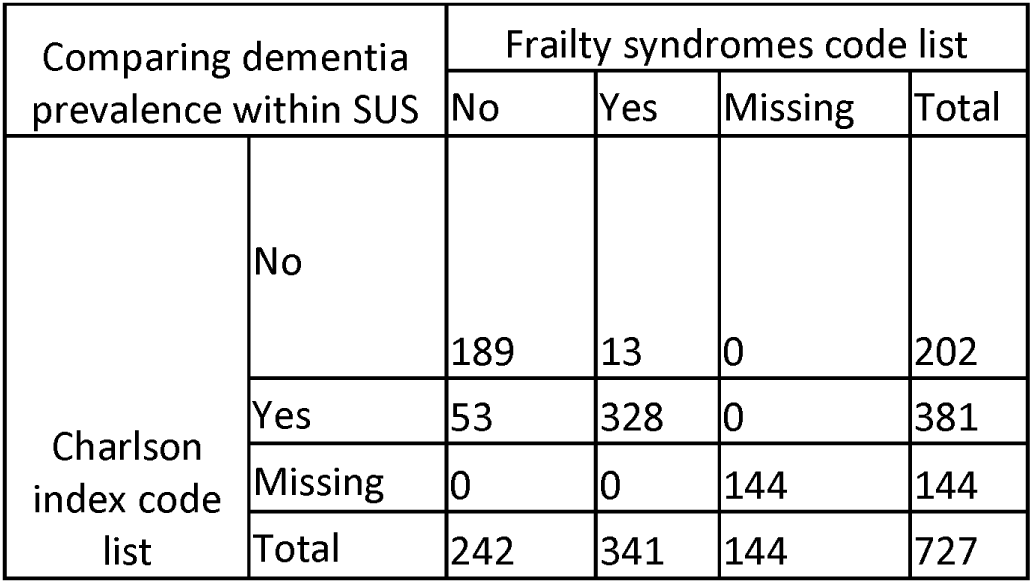
Table a) Comparison of residents identified to have dementia based on Charlson index code list and Frailty syndromes code list

| Comparing dementia prevalence within SUS |  | Frailty syndromes code list |  |  |  |
| --- | --- | --- | --- | --- | --- |
|  |  | No | Yes | Missing | Total |
| Charlson index code list | No | 189 | 13 | 0 | 202 |
|  | Yes | 53 | 328 | 0 | 381 |
|  | Missing | 0 | 0 | 144 | 144 |
|  | Total | 242 | 341 | 144 | 727 |

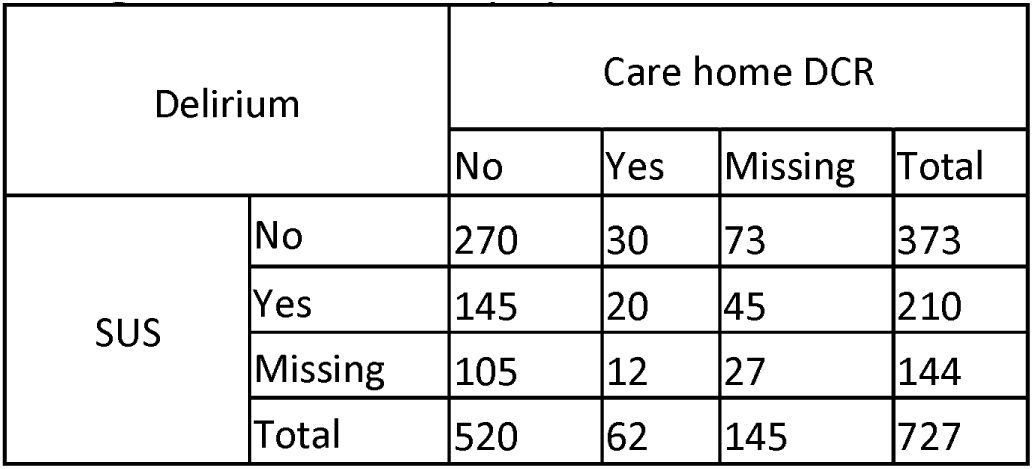
Table b) Comparison of residents identified to have delirium based on SUS data using Soong et al. List of frailty syndromes and care home DCR using I-AGED

| Delirium |  | Care home DCR |  |  |  |
| --- | --- | --- | --- | --- | --- |
|  |  | No | Yes | Missing | Total |
| SUS | No | 270 | 30 | 73 | 373 |
|  | Yes | 145 | 20 | 45 | 210 |
|  | Missing | 105 | 12 | 27 | 144 |
|  | Total | 520 | 62 | 145 | 727 |

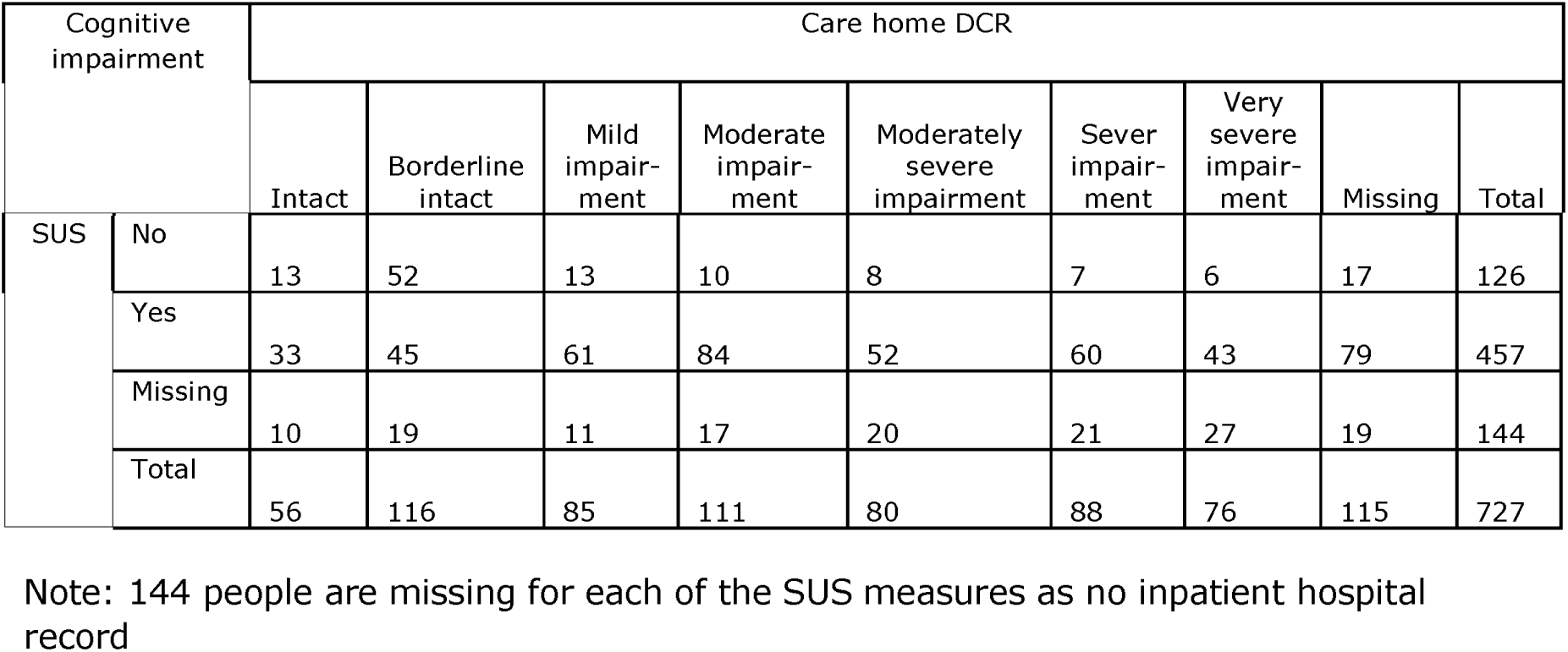
Table c) Comparison of residents identified to have cognitive impairment based on Soong et al. List of frailty syndromes and care home DCR, assess using Morris et al.

| Cognitive impairment |  | Care home DCR |  |  |  |  |  |  |  |  |
| --- | --- | --- | --- | --- | --- | --- | --- | --- | --- | --- |
|  |  | Intact | Borderline intact | Mild impairment | Moderate impairment | Moderately severe impairment | Sever impairment | Very severe impairment | Missing | Total |
| SUS | No | 13 | 52 | 13 | 10 | 8 | 7 | 6 | 17 | 126 |
|  | Yes | 33 | 45 | 61 | 84 | 52 | 60 | 43 | 79 | 457 |
|  | Missing | 10 | 19 | 11 | 17 | 20 | 21 | 27 | 19 | 144 |
|  | Total | 56 | 116 | 85 | 111 | 80 | 88 | 76 | 115 | 727 |

Note: 144 people are missing for each of the SUS measures as no inpatient hospital record

## APPENDIX 10 Complete final prototype MDS. Numbers **are** reported for 727 residents unless otherwise specified

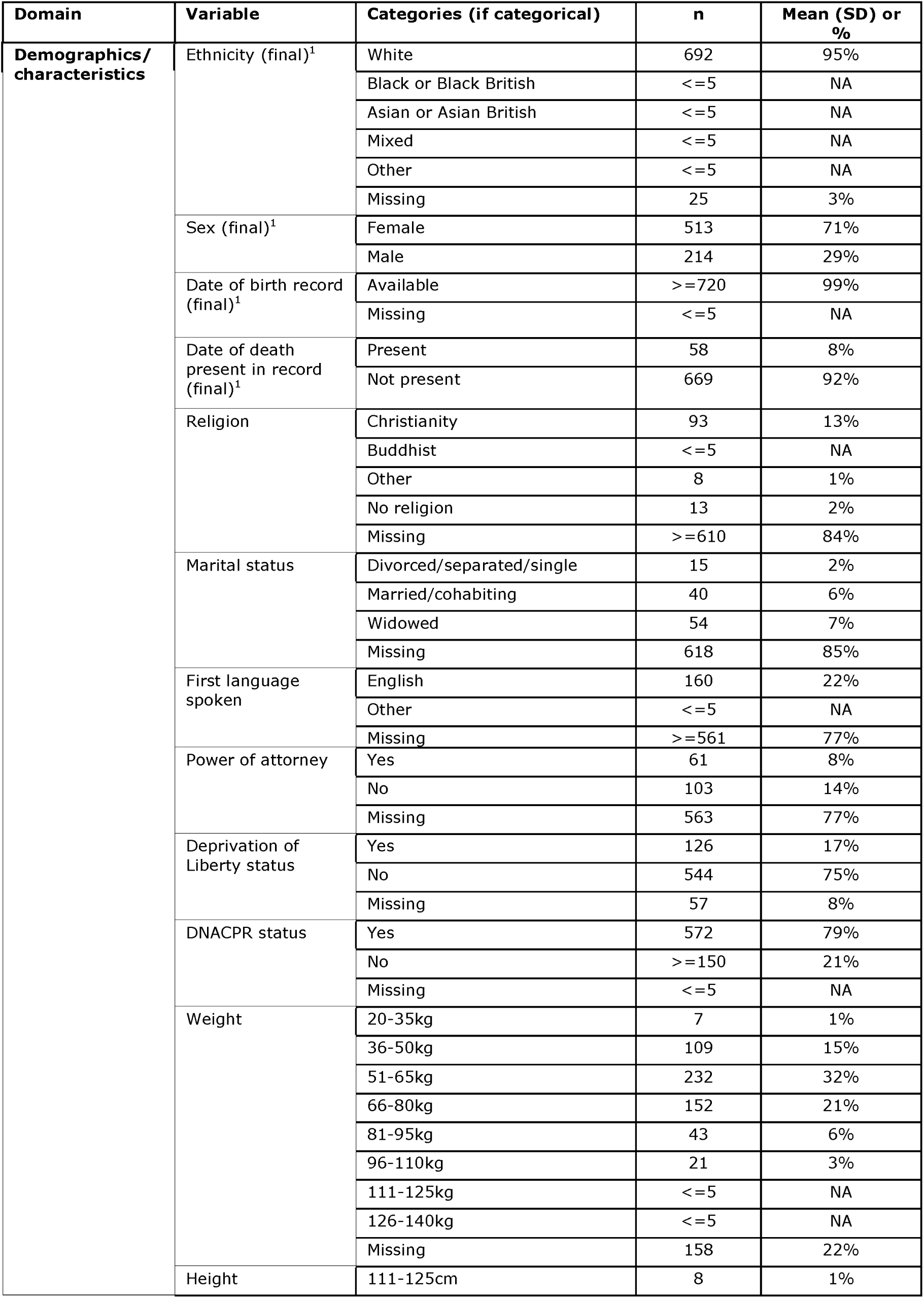
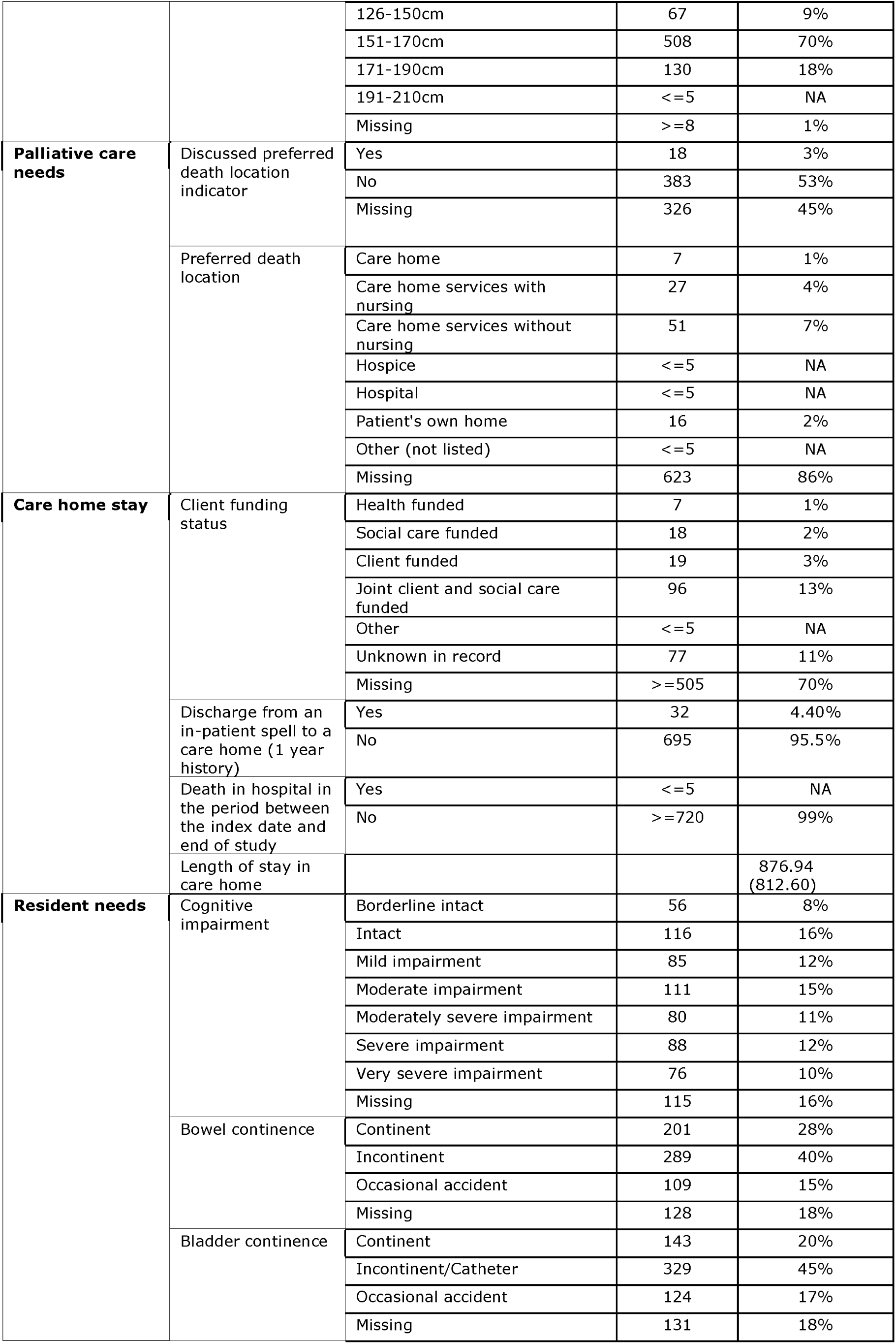
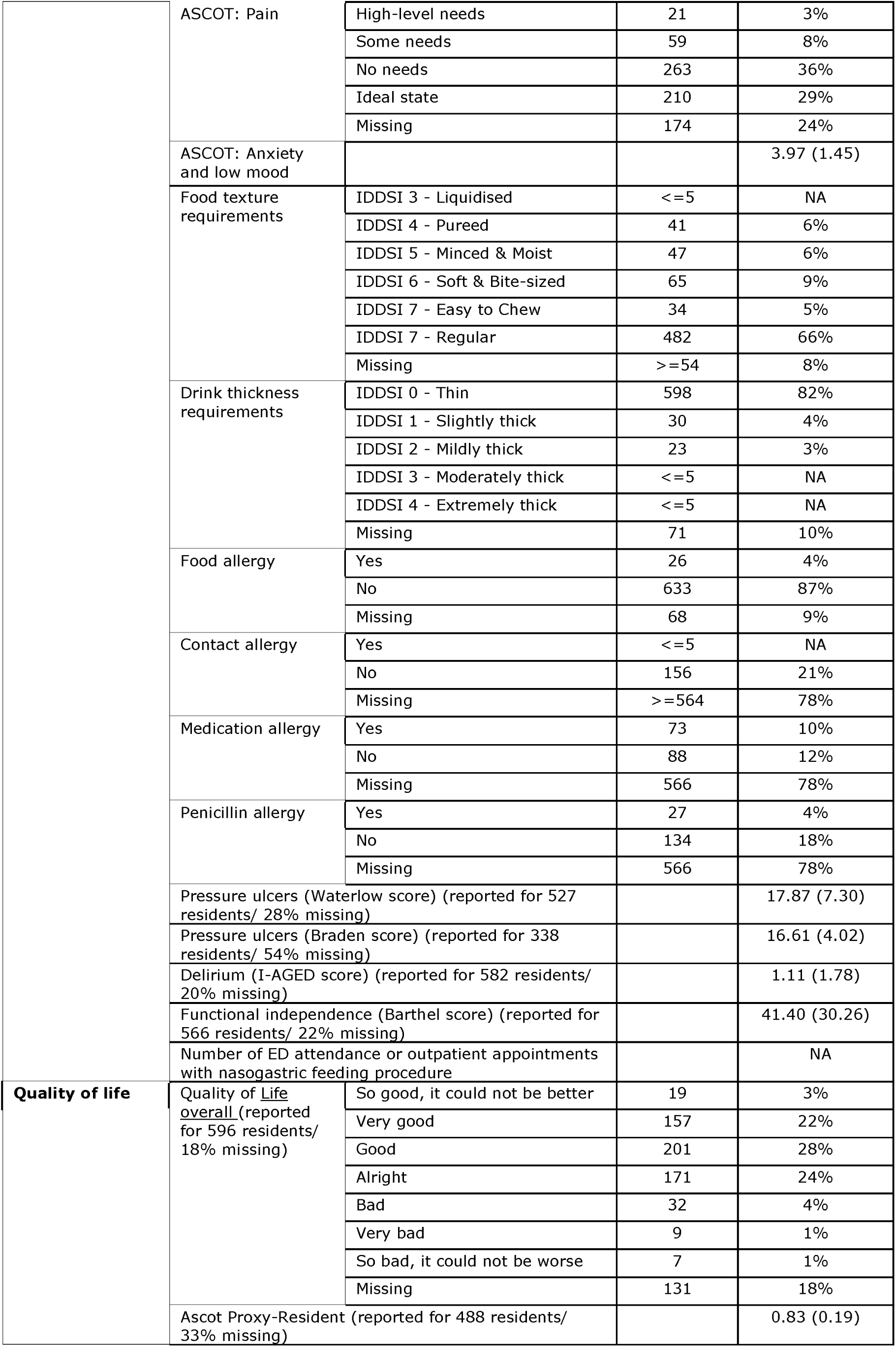
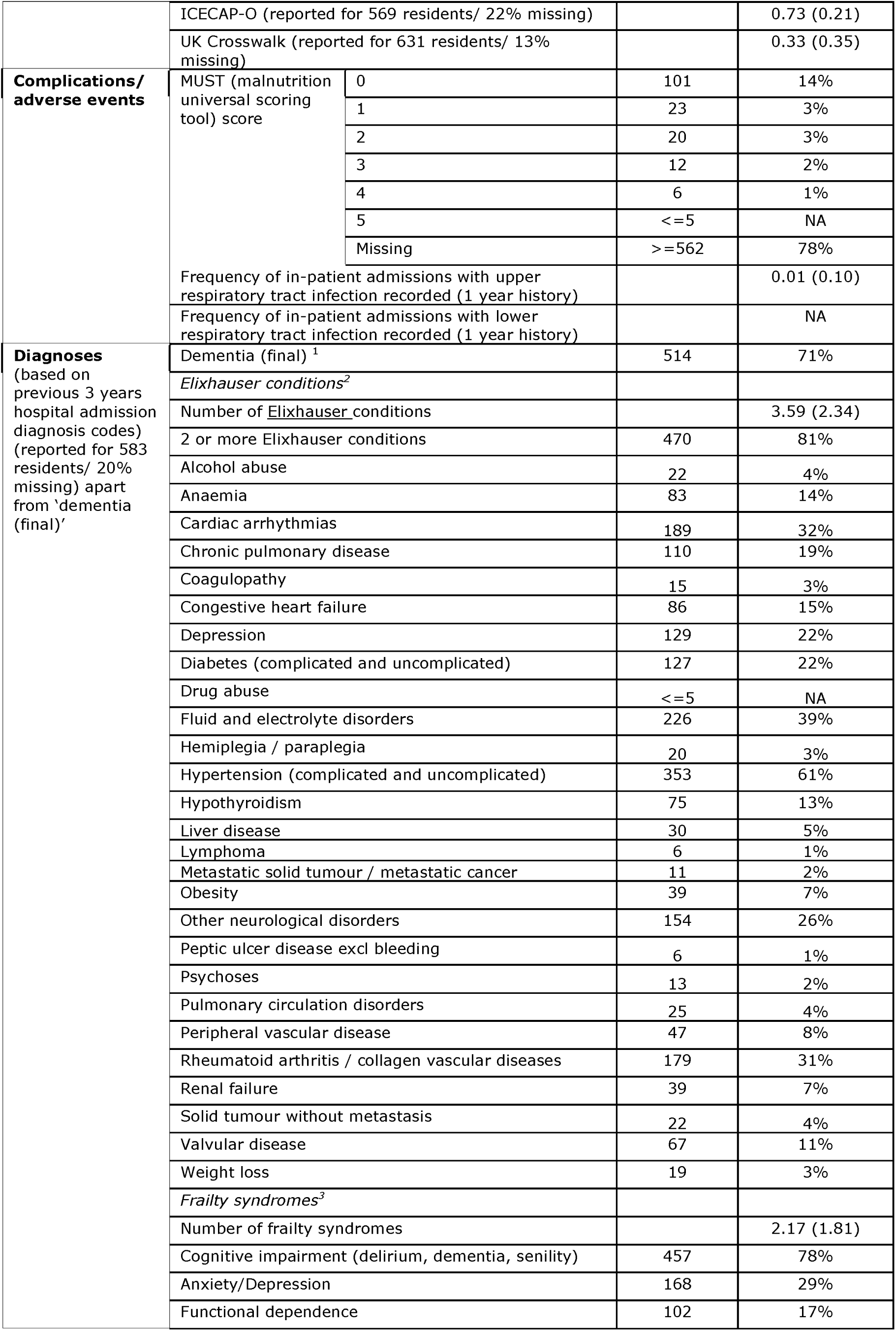
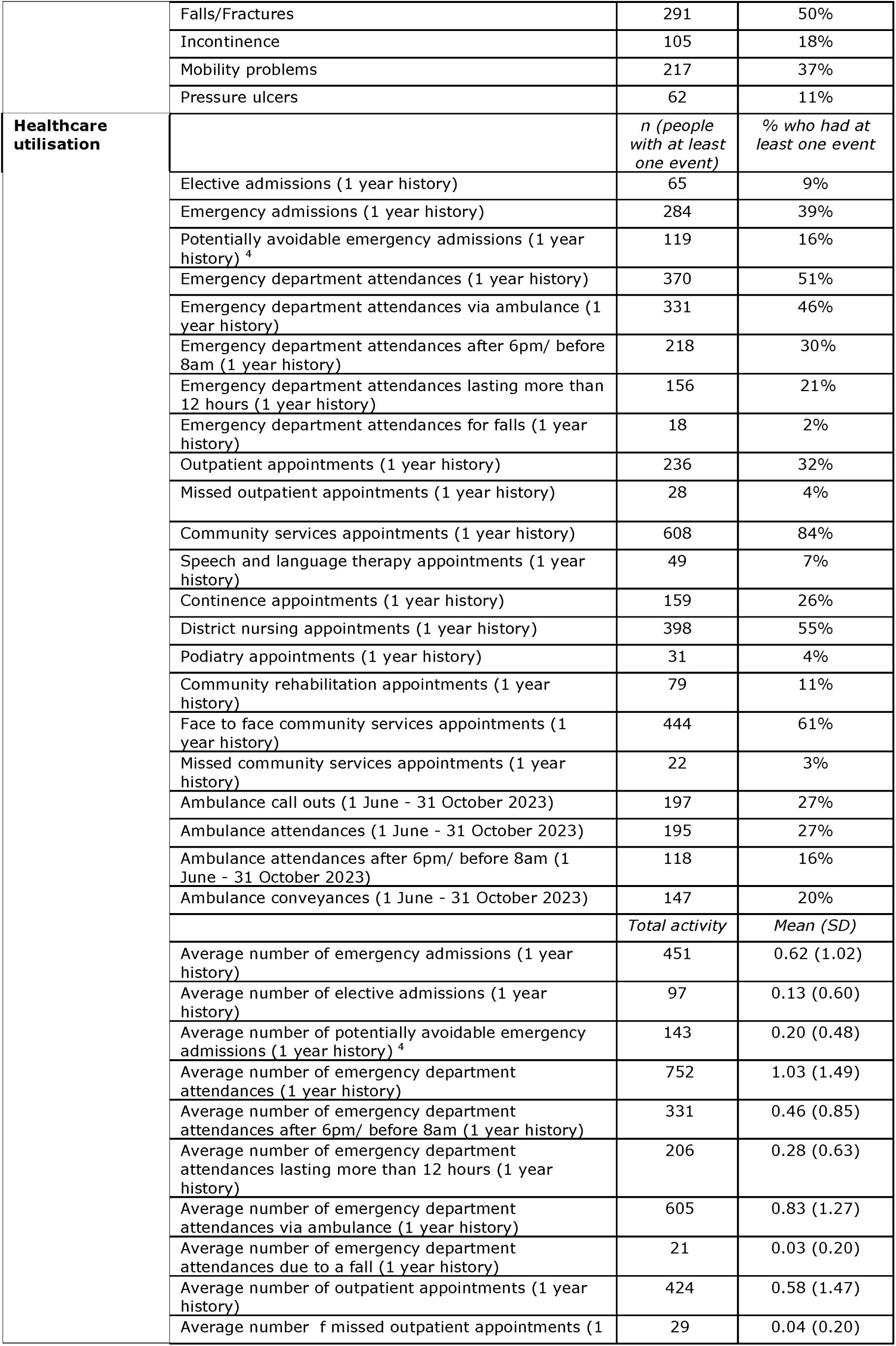
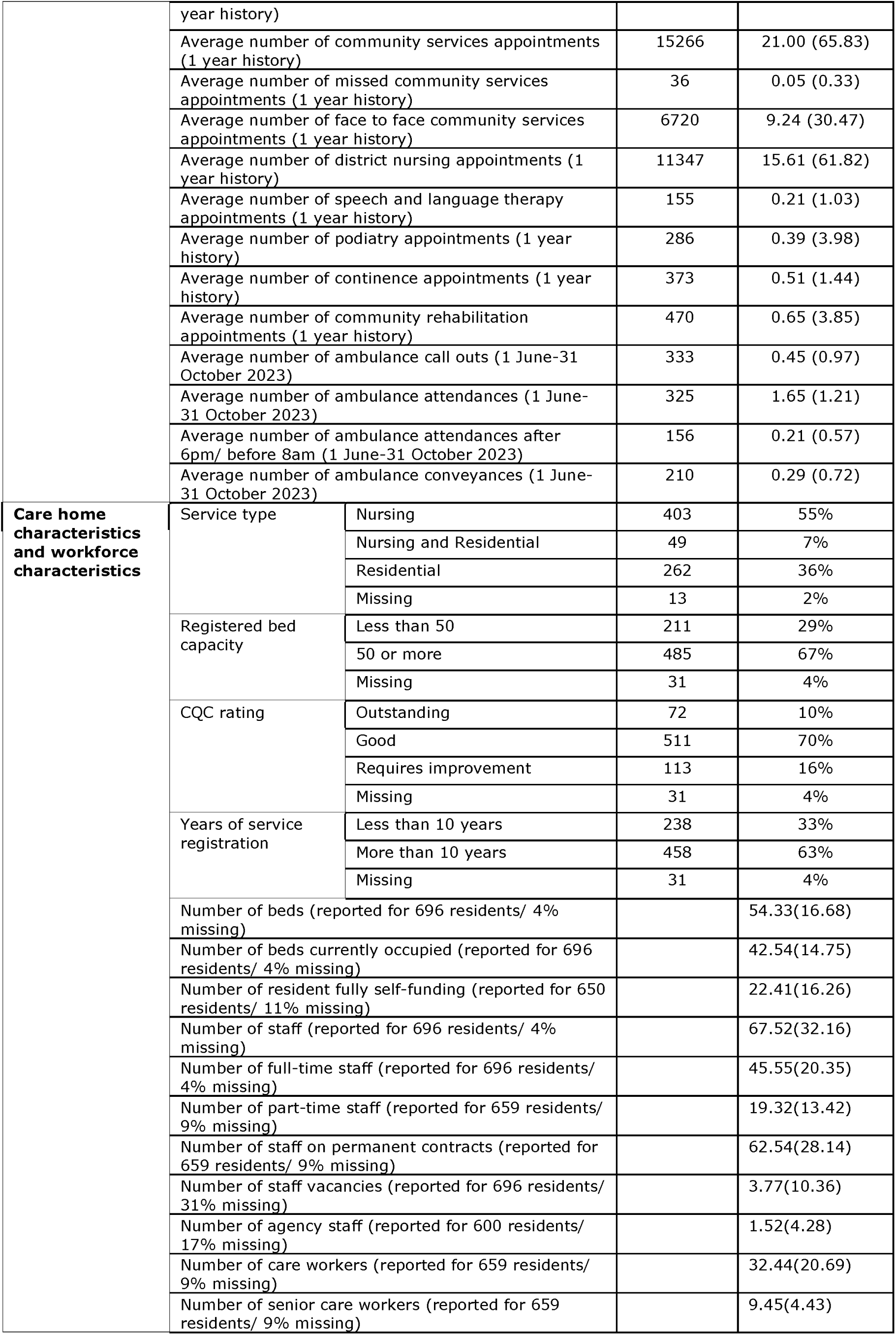
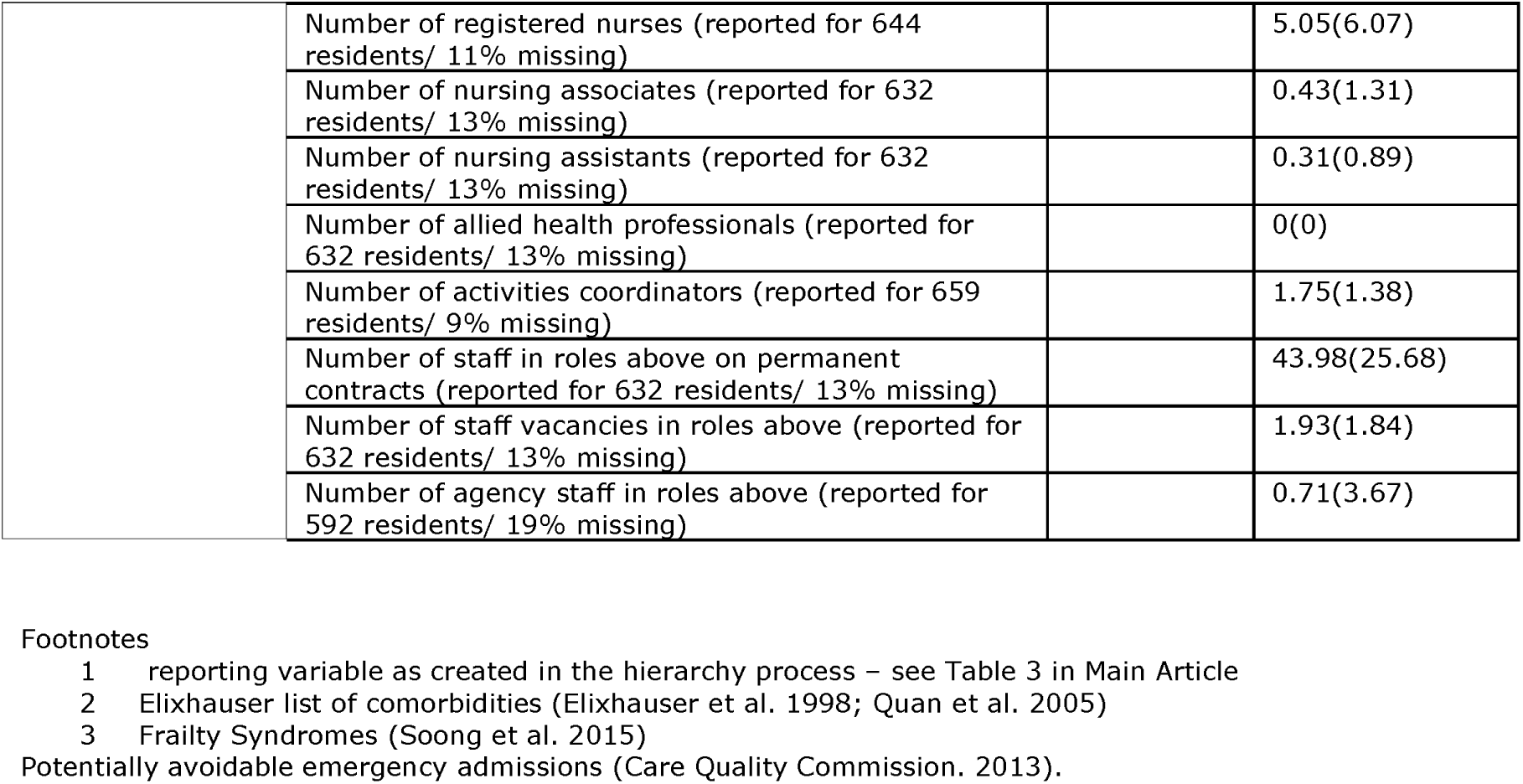

## APPENDIX 11 Using MDS data to understand ED attendance and ambulance activity for care home residents: a worked example

In the main report, we presented key variables from the MDS with a full description of the MDS in Appendix 8. However, a key benefit of an MDS is the ability to explore sub groups of residents. These tables are examples of analyses that could help understand whether there are differences in outcomes or activities in different subgroups. Tables below are examples of such subgroup analyses which were suggested by stakeholders as of interest. For example, in our sample population, activity was in general higher across both A&E and ambulance services for those in residential care homes, compared to nursing homes, which could be informative when commissioning local services.

Table a) ED attendances for year leading to index date 1. * where no mean reported as denominator <=5. Mean reports the mean of all those eligible e.g. can only have ED attendance via ambulance if you have had an ED attendance

| Variable | Total | Sex |  | Age |  |  |  | Nursing vs residential |  |  | Dementia |  |  | Deprivation<sup>1</sup> |  |  |  |  |  |
| --- | --- | --- | --- | --- | --- | --- | --- | --- | --- | --- | --- | --- | --- | --- | --- | --- | --- | --- | --- |
|  |  | Female | Male | <65 | 65-79 | >=80 | Missing | Nursing | Residential | Missing | No | Yes | Missing | Most deprived fifth | Second most deprived quintile | Middle fifth | Second least deprived fifth | Least deprived fifth | Missing |
| N | 727 | 513 | 214 | 17 | 125 | 486 | 99 | 452 | 262 | 13 | 191 | 514 | 22 | 81 | 158 | 72 | 119 | 198 | 99 |
| Mean (SD) |  |  |  |  |  |  |  |  |  |  |  |  |  |  |  |  |  |  |  |
| Number of A&E attendances | 1.03<br>(1.49) | 0.94<br>(1.44) | 1.25<br>(1.61) | 1.06<br>(1.34) | 1.05<br>(1.46) | 0.93<br>(1.41) | 1.51<br>(1.85) | 0.92<br>(1.44) | 1.21<br>(1.57) | 1.38<br>(1.50) | 0.92<br>(1.39) | 1.11<br>(1.55) | * | 0.95<br>(1.37) | 1.02<br>(1.40) | 1.04<br>(1.30) | 1.13<br>(1.63) | 0.79<br>(1.35) | 1.51<br>(1.85) |
| Number of out of hours A&E attendances | 0.46<br>(0.85) | 0.41<br>(0.77) | 0.57<br>(1.00) | 0.76<br>(1.25) | 0.50<br>(0.81) | 0.41<br>(0.81) | 0.56<br>(0.95) | 0.39<br>(0.82) | 0.56<br>(0.90) | * | 0.38<br>(0.81) | 0.50<br>(0.87) | * | 0.52<br>(1.01) | 0.47<br>(0.78) | 0.47<br>(0.87) | 0.48<br>(0.85) | 0.35<br>(0.75) | 0.56<br>(0.95) |
| Number of A&E attendances lasting >12 hours | 0.28<br>(0.63) | 0.25<br>(0.62) | 0.36<br>(0.65) | * | 0.35<br>(0.65) | 0.25<br>(0.59) | 0.36<br>(0.80) | 0.26<br>(0.58) | 0.32<br>(0.72) | * | 0.29<br>(0.71) | 0.29<br>(0.62) | * | 0.07<br>(0.26) | 0.34<br>(0.66) | 0.32<br>(0.60) | 0.21<br>(0.57) | 0.32<br>(0.66) | 0.36<br>(0.80) |
| Number of A&E attendances via ambulance | 0.83<br>(1.27) | 0.76<br>(1.22) | 1.00<br>(1.37) | 0.71<br>(1.16) | 0.82<br>(1.23) | 0.79<br>(1.27) | 1.09<br>(1.33) | 0.72<br>(1.22) | 1.02<br>(1.34) | 1.00<br>(1.15) | 0.76<br>(1.23) | 0.89<br>(1.30) | * | 0.85<br>(1.29) | 0.91<br>(1.26) | 0.90<br>(1.13) | 0.79<br>(1.40) | 0.63<br>(1.18) | 1.09<br>(1.33) |
| Number of A&E attendances for falls | 0.03<br>(0.20) | 0.02<br>(0.16) | 0.04<br>(0.26) | * | * | 0.02<br>(0.19) | 0.08<br>(0.31) | * | 0.06<br>(0.30) | * | * | 0.04<br>(0.22) | * | * | * | * | * | * | 0.08<br>(0.31) |

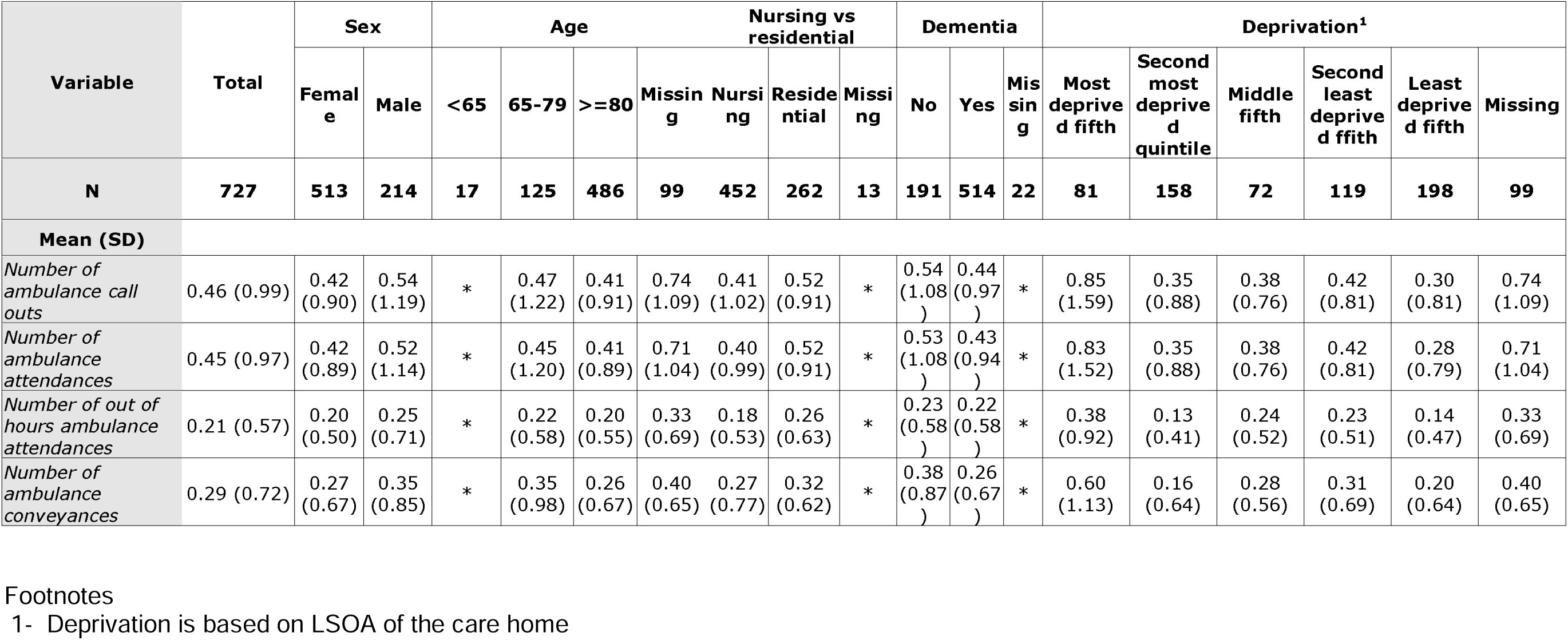
Table b) Ambulance activity for 5 month period from 1 June to 31 October. * where no mean reported as denominator <=5. Mean reports the mean of all those eligible e.g. can only have ED attendance via ambulance if you have had an ED attendance

| Variable | Total | Sex |  | Age |  |  | Nursing vs residential |  |  |  | Dementia |  |  | Deprivation<sup>1</sup> |  |  |  |  |  |
| --- | --- | --- | --- | --- | --- | --- | --- | --- | --- | --- | --- | --- | --- | --- | --- | --- | --- | --- | --- |
|  |  | Female | Male | <65 | 65-79 | >=80 | Missing | Nursing | Residential | Missing | No | Yes | Missing | Most deprived fifth | Second most deprived quintile | Middle fifth | Second least deprived fifth | Least deprived fifth | Missing |
| N | 727 | 513 | 214 | 17 | 125 | 486 | 99 | 452 | 262 | 13 | 191 | 514 | 22 | 81 | 158 | 72 | 119 | 198 | 99 |
| Mean (SD) |  |  |  |  |  |  |  |  |  |  |  |  |  |  |  |  |  |  |  |
| Number of ambulance call outs | 0.46 (0.99) | 0.42 (0.90) | 0.54 (1.19) | * | 0.47 (1.22) | 0.41 (0.91) | 0.74 (1.09) | 0.41 (1.02) | 0.52 (0.91) | * | 0.54 (1.08) | 0.44 (0.97) | * | 0.85 (1.59) | 0.35 (0.88) | 0.38 (0.76) | 0.42 (0.81) | 0.30 (0.81) | 0.74 (1.09) |
| Number of ambulance attendances | 0.45 (0.97) | 0.42 (0.89) | 0.52 (1.14) | * | 0.45 (1.20) | 0.41 (0.89) | 0.71 (1.04) | 0.40 (0.99) | 0.52 (0.91) | * | 0.53 (1.08) | 0.43 (0.94) | * | 0.83 (1.52) | 0.35 (0.88) | 0.38 (0.76) | 0.42 (0.81) | 0.28 (0.79) | 0.71 (1.04) |
| Number of out of hours ambulance attendances | 0.21 (0.57) | 0.20 (0.50) | 0.25 (0.71) | * | 0.22 (0.58) | 0.20 (0.55) | 0.33 (0.69) | 0.18 (0.53) | 0.26 (0.63) | * | 0.23 (0.58) | 0.22 (0.58) | * | 0.38 (0.92) | 0.13 (0.41) | 0.24 (0.52) | 0.23 (0.51) | 0.14 (0.47) | 0.33 (0.69) |
| Number of ambulance conveyances | 0.29 (0.72) | 0.27 (0.67) | 0.35 (0.85) | * | 0.35 (0.98) | 0.26 (0.67) | 0.40 (0.65) | 0.27 (0.77) | 0.32 (0.62) | * | 0.38 (0.87) | 0.26 (0.67) | * | 0.60 (1.13) | 0.16 (0.64) | 0.28 (0.56) | 0.31 (0.69) | 0.20 (0.64) | 0.40 (0.65) |

